## Supplementary materials for "Cognitive Performance in Relation to Systemic and Brain Iron at Perimenopause"

### 1. Methodological details

#### *1.1. Face/name associative memory task (FNAM)*

The FNAM measures episodic memory for the association of faces with names and putative occupations and has been shown to show some discriminative sensitivity to the presence of mild cognitive impairment [1–3]. The version of the FNAM used in the present study was based on that used by Rubiño and colleagues [4]. The FNAM consisted of three phases: a learning phase followed by an initial recall phase, and a final delayed recall phase, which occurred approximately two hours after the initial learning and recall phases (at the end of the testing session).

In the initial learning phase, participants were first shown photographs of a set of 24 women with each image presented for 2 sec. Facial images used as stimuli in the face name associative memory task were obtained from publicly available websites and were resized and converted to grayscale and were arbitrarily and randomly paired with a set of names and a set of occupations. The facial images subtended 4.8° of visual angle, both horizontally and vertically. Order of presentation in all phases was randomized across participants. Note that the algorithm that evaluates submissions to `medrxiv` ignored the description of the images and the assignment of names and occupations and erroneously determined that the example images contained personally identifying information and requested that we remove the example. Consequently the example images and pairings can be obtained on request to the corresponding author.

Participants self-initiated the sequence of presentations in each of the learning phases. After the initial presentation of all the faces, each face was shown again, now with a name directly below the photograph (Supplementary Figure **??**b). Participants were instructed to study each face and name pairing for 2 sec. After studying the faces and names, participants were then tested on their memory for the association of faces and names. On each test trial, participants were presented with a studied face along with two names, below and to the left and right of the face; one of the names was the originally studied name and one was a novel lure (Supplementary Figure **??**c). Participants pressed the z key if they thought the correct name was presented below the photograph on the left side of the screen and pressed the m key if the correct name was on the right side. Participants were given 2.5 seconds to respond to each photograph, and feedback was given for 1.5 sec on their choice after a 1 sec delay.

In the next stage, participants learned the occupations of each of the 24 women they had previously seen. Participants self-initiated the set of presentations. Each face appeared on the screen for 2 sec with an occupation label (Supplementary Figure **??**d) listed directly below the face. Following the presentation of all the face-occupation pairings, participants were tested on their immediate memory for those associations. The test trials followed the same procedure used to test the initial face-name associations (Supplementary Figure **??**e). After all the other behavioral tests were completed, the final (delayed, after approximately 2 hours) test of memory for the face-name and face-occupation associations was performed using the same procedures as were used for the initial tests.

#### *1.2. Probabilistic selection task (PST)*

The PST measures positive and negative reinforcement learning, which can be affected by variations in levels of dopamine [5,6]. The stimuli in the PST were characters from the Nepalese Devanagari script presented in pairs with each member of the pair subtending 4.8° horizontally and vertically, and each stimulus was offset 75 pixels horizontally from the center of the screen.

The PST involved two phases: a training phase and a testing phase. In the training phase, participants were presented with three pairs of stimuli (see Supplementary Figure S1) and needed to learn which member of each pair was rewarded more frequently. One member of each pair was rewarded more frequently than the other, and the differential reward rate was probabilistic. For the first pair of stimuli (the AB pair), A was rewarded 80% of the time while B was rewarded 20% of the time. For the second pair (the CD pair), the relative rates of reward were 70% vs. 30%, and for the third pair (the EF pair), the relative rates of reward were 60% vs. 40%. These pairings were presented in blocks, with each block containing three presentations of each pairing in random order. Each trial in each block was self-initiated, followed by the presentation of a fixation cross with an exposure duration drawn from an exponential distribution with a mean of 750 ms, censored at 500 and 1000 ms. The test pairing for the trial was then presented with left/right location of each member of the pair determined randomly on each trial. The pair was presented for up to 3 sec or until the participant made a choice. Participants were instructed to choose the rewarded member of the pair using the z key for the member on the left and the m key for the right. Visual feedback ('correct' or 'incorrect') was presented for 1.5 seconds after a 0.7 sec delay. Participants exited the training phase if either they met criterion levels of preference for the three pairs (choosing A 65%

of the time in AB pairings, choosing C 60% of the time in CD pairings, and choosing E 50% of the time in EF pairings) in a given block, or they completed three blocks of training trials.

The testing phase involved the presentation of all possible pairings of the six stimuli and tested the choice preferences learned in the training phase from two perspectives. The first was with respect to the extent to which learning to choose stimulus A (the most rewarded stimulus) and learning to avoid stimulus B (the least rewarded stimulus) generalized to other pairings. The other was with respect to the amount of conflict present in the tested choice. This took two forms: conditions of high conflict, involving a choice between either two frequently rewarded stimuli (A with C or E, C with E) or two rarely rewarded stimuli (B with D or F, D with. F); and conditions of low conflict, involving a choice between a frequently rewarded and an infrequently rewarded stimulus (A with D or F, B with C or E). Each of the six possible pairings was repeated 15 times, with the order of presentation determined randomly. The trial events in the testing phase were identical to those in the training phase, with the exception that no feedback was provided.

#### *1.3. Rule-based category learning task (RBCL)*

The RBCL is drawn from a literature on how the learning of different category structures is hypothesized to require the involvement of two distinct forms of learning and memory [7–10]. One of these is procedural learning, which is dependent on implicit learning of stimulus response associations, and the second is declarative learning, which is dependent on learning explicit, verbalizable rules. There are data suggesting the involvement of DA in both types of learning, with recent work [11] suggesting that DA functions in declarative learning with respect to the maintenance and switching of response rules and in procedural learning as a training signal.

In the RBCL, participants were presented with Gabor patches varying on two dimensions: spatial frequency and orientation. Stimuli subtended 2.8° both horizontally and vertically and were centered on the screen.Participants were required to learn the stimuli and place them in one of four categories, labeled A, B, C, and D. A total of 100 stimuli were generated for each category by drawing from Gaussian distributions parametrized for spatial frequency and orientation; see Supplementary Table S1 for these means and standard deviations of these distributions and see Supplementary Figure S2 for example stimuli from each category. The rule for determining category was a simple verbalizable rule, such that if orientation and spatial frequency were both low, the stimulus belonged to category A, and if orientation and spatial frequency were both high, the stimulus belonged to category D, etc..

At the beginning of the task, participants were shown examples of each category without any information about the rule that determined its category membership. Participants self-initiated each trial using the space bar in response to a circle that appeared on the screen. A fixation cross with a duration drawn from an exponential distribution with a mean of 750 ms, censored at 500 and 1000 ms. The test stimulus was presented until a response was made or until 2 s elapsed. Visual feedback was presented for 1.5 s following the participant's choice. The message "…take a short break …" appeared after every 100 trials. Participants then continued until 400 trials were completed, with stimuli from each of the categories presented in random order 100 times. At the end of the task participants were asked, "For the task you

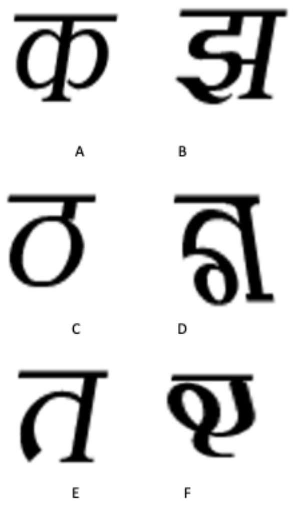

**Figure S1.** Examples of the presentation of stimuli during the training and test phases of the PST. See text for details of the pairings during the training phase.

**Table S1.** Parameters for the Gaussian distributions used to generate stimuli for the RBCL.

| | Orientation (degrees) | | Spatial frequency (cycles/100 pixels) | |
|---|---|---|---|---|
| Category | $\mu$ | $\sigma$ | $\mu$ | $\sigma$ |
| A | 54 | 8 | 4 | 1 |
| B | 54 | 8 | 6 | 1 |
| C | 76 | 8 | 4 | 1 |
| D | 76 | 8 | 6 | 1 |

just finished, you should have been making your decisions based on two dimensions. What do you think those two dimensions were? How were you making your decisions?" to check if they could verbalize the rules from the task.

#### *1.4. Visuospatial working memory task (VSWM)*

The VSWM was included on the basis of a set of findings [12–14] suggesting that variations in performance on tests of this form of working memory can be related to variations in DA status as well as age [15]. In the current study, visuospatial working memory is dependent on both attention and memory.

In the practice phase, two types of trials were introduced. In all practice trials, after a small circle appeared on the screen, participants were instructed to press the space bar to initiate the trial. A fixation cross with an exposure duration drawn from an exponential distribution with a mean of 750 ms, censored at 500 and 1000 ms, appeared on the screen. This was followed by a study display comprised of 16 white squares arranged in a circle. Three targets (red circles) randomly appeared in each of the 16 squares. Participants were instructed to remember the locations of the three targets. This study display was presented for 1 s followed by a 2 s blank display. The test display then followed, comprised of the 16 white squares (arranged in a circle) with a question mark appearing in one of the squares, prompting the participant to indicate whether a target was present in that square in the study display. Participants gave a positive response with the index finger of their dominant hand and a negative response with the index finger of their non-dominant hand. Participants were given up to 3 sec to respond before the next trial began, and feedback was not presented. Five practice trials were completed in the first set.

In the second set of practice trials, distractors (blue circles) were included along with the targets. The second set of practice trials followed the same process as the first set. Sixteen white squares arranged in a circle appeared on the screen. Three targets (red circles) and two distractors (blue circles) randomly appeared in each of the 16 squares. Participants were instructed to remember the locations of the three targets and to ignore the locations of the distractors. This study display was presented for 1 s followed by a 2 s blank display. The test display then followed, comprised of the 16 white

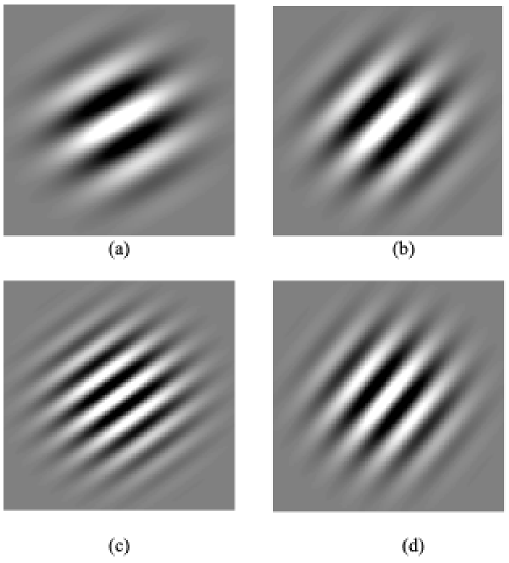

**Figure S2.** Examples of the stimuli from the four categories used in the RBCL: (a) low spatial frequency, low orientation; (b) low spatial frequency, high orientation; (c) high spatial frequency, low orientation; (d) high spatial frequency, high orientation.

squares with a question mark appearing in one of the squares, prompting the participant to indicate whether or not a target was present in that square in the study display. Participants gave a positive response with the index finger of their dominant hand and a negative response with the index finger of their non-dominant hand. Participants were given up to 3 sec to respond before the next trial began, and feedback was not given.

Each trial in the testing phase of the VSWM was self-initiated. After a small circle appeared on the screen, participants were instructed to press the space bar to initiate the trial. A fixation cross with an exposure duration drawn from an exponential distribution with a mean of 750 ms, censored at 500 and 1000 ms, appeared on the screen. This was followed by a study display comprised of 16 white squares arranged in a circle appeared on the screen. A randomly selected set of either three or five squares contained targets (red circles) while another randomly selected set of either zero or two additional squares contained distractors (blue circles). Participants were instructed to remember the locations of the red circles and to ignore the locations of the blue circles (see Supplementary Figure S3 for an example of a study display). This study display was presented for 1 s followed by a 2 s blank display. The test display then followed, comprised of the 16 white squares (arranged in a circle) with a question mark appearing in one of the squares, prompting the participant to indicate whether a target was present in that square in the study display. Participants gave a positive response with the index finger of their dominant hand and a negative response with the index finger of their non-dominant hand. Participants were given up to 3 sec to respond before the next trial began, and feedback was not presented. A total of 96 trials were presented, with half of the trials probing a studied location and half probing an unstudied location.

### 2. EEG acquisition, pre-processing, and feature extraction

#### *2.1. Acquisition*

EG data were collected using high-density (128 channel) electrode nets (Magstim/EGI, Eugene OR). Data were acquired using a 128-channel Net Amps 300 amplifier and were digitized at a sampling rate of 1 kHz. Impedances were kept at or below 50 $K\Omega$ during testing. Data were collected with online filters set at 0.1 Hz (highpass), 70 Hz (lowpass), and 60 Hz (notch).

#### *2.2. Pre-processing*

EEG data from all of the tasks and the resting data were pre-processed up to the point of feature extraction using the same set of steps. All pre-processing and feature extraction were done using the EEGLAB toolbox [16] for Matlab (Mathworks, Natick MA) following a recently published pipeline [17]. Data were first filtered using a high-pass filter at 0.5 Hz, a low-pass filter at 90 Hz, and a notch filter at 58-62 Hz. All data from before the start of the task and after the end of the task were then removed, bad channels identified by visual inspection were removed, and then bad segments and any additional bad channels were removed using the function `pop_clean_rawdata` in EEGLAB. Data were then re-referenced to the mean. An independent component analysis was then done to identify artifacts such as muscle, eye, and cardiac artifacts [18]. The EEGLAB functions `pop_iclabel` and `pop_icflag` were then used to flag and remove components identified as artifacts. Missing channels were then interpolated.

#### *2.3. Feature extraction: FNAM*

The features extracted for the FNAM data were based on those in Guo et al. 2005 and Mitchell et al. 2016. Pre-processed data were low-pass filtered at 30 Hz and data were restricted to those recorded at electrodes 11, 129, 62, and 75, corresponding to Fz, Cz, Pz, and Oz, for the testing phase of the task. Data were then epoched from 200 ms preceding

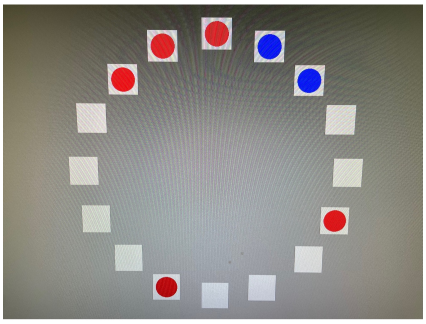

**Figure S3.** Examples of a study display in the VSWM, containing five targets (red circles) and two distractors (blue circles).

to 2500 ms following the onset of the test stimulus and baseline corrected. Average waveforms for remembered and forgotten face/name and face/occupation associations were then calculated for each participant. From these average waveforms, mean amplitudes were then calculated for three time periods following the onset of the test stimulus: 200-400 ms (positive amplitudes), 400-600 ms (negative amplitudes), and 600-800 ms (positive amplitudes).

#### 2.4. Feature extraction: PST

Response-locked and feedback-locked features for the PST data were based on those in Frank et al. 2005, Larson et al. 2015, and Schmid et al. 2018. The response-locked features were error-related negativity (ERN) and positivity (Pe), and correct-related negativity (CRN). The feedback-locked features were the feedback-related negativity (FRN) for correct and incorrect responses. All features were extracted from the data from the test phase of the task. Pre-processed data were low-pass filtered at 15 Hz and then were epoched from 200 ms prior to either the response or the feedback to 500 ms after the response or 800 ms after the feedback. Data for the ERN and CRN were drawn from a cluster of electrodes surrounding Cz (4, 5, 11, 12, 16, and 19, following [21]). Data for the Pe were drawn from a separate set of electrodes (4, 55, 61, 62, 78, and 79, following [22]). Data for the FRN were drawn from a set of three electrodes (6, 7, and 106, following [23]). Average waveforms were calculated for each participant and baseline corrected.

The ERN was calculated as the difference between the maximum negative amplitude within 200 ms of the onset of the stimulus and the preceding maximum positive amplitude. The Pe was calculated as the difference between the maximum positive amplitude within 200 ms of the onset of the stimulus and the preceding maximum negative amplitude. The CRN was calculated as the difference between the maximum negative amplitude within 400 ms of the onset of the stimulus and the preceding maximum positive amplitude. The FRN for both correct and incorrect responses was calculated as the difference between the maximum negative amplitude in the interval running from 100-300 ms from the onset of the feedback and the maximum positive amplitude preceding it.

#### 2.5. Feature extraction: RBCL

Stimulus- and feedback-locked features for the RBCL data were based on those in Rabi et al. 2018. Stimulus-locked features were obtained from electrodes 58, 59, 62, 91, and 96, while feedback-locked features were obtained from electrodes 11, 24, 36, 54, 62, 79, 104, and 124. Pre-processed data were low-pass filtered at 30 Hz and then were epoched from 200 ms prior to 800 ms after the onset of either the stimulus or the feedback. The data were averaged separately for correct and incorrect responses for each participant and then baseline corrected. The stimulus-locked P300 was calculated as the mean amplitude in the interval running from 300-600 ms after the onset of the stimulus. The feedback-locked late positive slow wave was calculated as the mean amplitude in the interval running from 300 to 650 ms following the onset of the feedback.

#### 2.6. Feature extraction: VSWM

Features for the VSWM data were based on those in Knott et al. 2004. Pre-processed data were low-pass filtered at 12 Hz, the data for correct and incorrect responses were epoched from 200 ms before to 1000 ms after the onset of the stimulus, averaged for each participant, and then baseline corrected. The P300 for each trial type was calculated as the maximum positive amplitude in the interval running from 250 to 800 ms following the onset of the stimulus in three electrodes (11, 55, and 62).

#### 2.7. Blink rates

Task-related and spontaneous blink rates were extracted from each of the tasks and the resting period using the BLINKER plug-in for EEGLAB [25].

### 3. MRI estimates of regional iron deposits

Our approach to obtaining estimates of regional brain iron deposits was based on that used in [26–28]. Specifically, we determined, for each participant across all ROIs, the intensity value that represented the lowest fifth percentile and isolated those pixels in each slice. We did this to ensure that we had the highest likelihood of isolating iron. We then averaged within and then across slices in each ROI in each hemisphere for each participating. These were the data that were subjected to analyses.

**Table S2.** Descriptive statistics for the behavioral data from the FNAM.

| Task | DV | Retention interval | M | SE | min | max |
|---|---|---|---|---|---|---|
| Face/name | P(C) | Immediate | 0.81 | 0.02 | 0.58 | 1.00 |
| | | Delayed | 0.74 | 0.03 | 0.4 | 0.92 |
| | RT (ms) | Immediate | 1402 | 57 | 998 | 1864 |
| | | Delayed | 1451 | 47 | 1037 | 1837 |
| Face/Occupation | P(C) | Immediate | 0.86 | 0.02 | 0.46 | 1.00 |
| | | Delayed | 0.85 | 0.02 | 0.46 | 0.96 |
| | RT (ms) | Immediate | 1418 | 46 | 813 | 1981 |
| | | Delayed | 1394 | 47 | 1141 | 1788 |
| Delayed recognition | Hit Rate | | 0.62 | 0.04 | 0.08 | 0.96 |
| | False alarm rate | | 0.14 | 0.19 | 0.04 | 0.33 |
| | $d'$ | | 1.5 | 0.2 | -0.72 | 2.91 |
| | $c$ | | 0.41 | 0.06 | -0.29 | 1.18 |
| Old items, correct responses | RT (ms) | | 1351 | 59 | 934 | 1981 |
| New items, correct responses | RT (ms) | | 1593 | 59 | 1115 | 2184 |

**Table S3.** Repeated-measures analyses of the behavioral data from the FNAM.

| | Factor | F | p |
|---|---|---|---|
| Accuracy, P(C) | Test type (TT) | 11.00 | 0.003 |
| | Retention interval (RI) | 3.72 | 0.066 |
| | TT x RI | 2.05 | 0.166 |
| RT, correct choices, ms | Test type (TT) | 0.26 | 0.613 |
| | Retention interval (RI) | 0.09 | 0.764 |
| | TT x RI | 0.86 | 0.363 |
| RT, Recognition | Old vs. New Items | 24.31 | < 0.001 |

### 4. Results

#### *4.1. FNAM*

##### 4.1.1. Behavioral data

Descriptive statistics for the behavioral data are presented in Supplementary Table S2. The behavioral data (accuracy and RT) were analyzed using 2 (test type: face/name, face/occupation) x 2 (retention interval: immediate, delayed) repeated measures mixed models analyses of variance (ANOVAs), with both factors manipulated within participants and participants as the random factor. Supplementary Table S3 presents the results of these analyses. Cells highlighted in gray indicate statistically significant results. In this case, the only significant effect was a main effect for test type in overall accuracy, with participants being more accurate in recalling face/occupation associations (0.86) than in recalling face/name associations (0.77).

##### 4.1.2. Correlations between behavior and iron status

The correlations between the behavioral variables in the FNAM are presented in Supplementary Tables S4 and S5.

**Table S4.** Correlations between the behavioral variables in the FNAM and the iron status biomarkers (1). Cells highlighted in gray indicate statistically significant results after controlling for the false discovery rate. Note: pctl = percentile.

| | Hb | | sFt | | sFt pctl | | Age | |
|---|---|---|---|---|---|---|---|---|
| Variable | *r* | *p* | *r* | *p* | *r* | *p* | *r* | *p* |
| P(C), immediate face/name | 0.25 | 0.255 | 0.30 | 0.165 | 0.46 | **0.029** | -0.41 | 0.052 |
| P(C), delayed face/name | 0.35 | 0.102 | 0.47 | **0.022** | 0.44 | 0.036 | -0.10 | 0.644 |
| P(C), immediate face/occupation | -0.07 | 0.763 | -0.19 | 0.395 | -0.16 | 0.456 | -0.04 | 0.858 |
| P(C), delayed face/occupation | 0.04 | 0.867 | 0.01 | 0.660 | 0.14 | 0.515 | -0.33 | 0.126 |
| Hit rate, delayed recognition | 0.46 | **0.026** | 0.48 | **0.020** | 0.48 | **0.022** | -0.15 | 0.493 |
| False alarm rate, delayed recognition | -0.09 | 0.693 | -0.32 | 0.139 | -0.23 | 0.284 | -0.02 | 0.929 |
| $d'$, delayed recognition | 0.34 | 0.112 | 0.49 | **0.016** | 0.46 | **0.026** | -0.11 | 0.626 |
| $c$, delayed recognition | -0.47 | **0.022** | -0.33 | 0.121 | -0.37 | 0.080 | 0.17 | 0.447 |
| RT, immediate face/name | -0.15 | 0.498 | -0.43 | 0.038 | -0.47 | **0.023** | 0.28 | 0.190 |
| RT, delayed face/name | 0.20 | 0.370 | -0.51 | **0.013** | -0.59 | **0.003** | 0.11 | 0.622 |
| RT, immediate face/occupation | 0.08 | 0.716 | -0.39 | 0.066 | -0.43 | 0.042 | 0.26 | 0.231 |
| RT, delayed face/occupation | -0.48 | **0.020** | -0.08 | 0.710 | -0.20 | 0.366 | 0.26 | 0.237 |
| RT, Old items | 0.12 | 0.573 | -0.52 | **0.012** | -0.56 | **0.006** | 0.04 | 0.839 |
| RT, New items | 0.09 | 0.695 | -0.49 | **0.018** | -0.60 | **0.002** | 0.19 | 0.395 |

##### 4.1.3. EEG data

Group average wave forms for remembered and forgotten face/name and face/occupation associations at each testing time and at each of the four electrodes are presented in Supplementary Figures S4 and S5. Amplitudes in each of the three time intervals at each of two retention intervals for the two types of association were analyzed using separate 4 (electrode: Fz, Cz, Pz, Oz) $\times$ 2 (recall status: remembered, forgotten) repeated measures mixed models with electrode and recall status as fixed factors and participants as random factors. The results of these analyses are presented in Supplementary Table S6.

**Table S5.** Correlations between the behavioral variables in the FNAM and the iron status biomarkers (2). Cells highlighted in gray indicate statistically significant results after controlling for the false discovery rate.

| | WBC | | RBC | | HCT | | MCV | | MCH | | MCHC | | RDW | |
|---|---|---|---|---|---|---|---|---|---|---|---|---|---|---|
| Variable | r | p | r | p | r | p | r | p | r | p | r | p | r | p |
| P(C), immediate face/name | -0.09 | 0.692 | 0.09 | 0.682 | 0.22 | 0.309 | 0.12 | 0.570 | 0.18 | 0.403 | 0.16 | 0.477 | -0.52 | **0.011** |
| P(C), delayed face/name | -0.07 | 0.739 | 0.03 | 0.883 | 0.17 | 0.430 | 0.16 | 0.479 | 0.39 | 0.068 | 0.53 | **0.009** | -0.45 | 0.030 |
| P(C), immediate face/occupation | 0.03 | 0.885 | 0.01 | 0.974 | 0.03 | 0.894 | 0.02 | 0.925 | -0.10 | 0.654 | -0.24 | 0.278 | 0.10 | 0.660 |
| P(C), delayed face/occupation | 0.01 | 0.957 | 0.00 | 0.986 | -0.01 | 0.955 | -0.02 | 0.924 | 0.04 | 0.861 | 0.13 | 0.569 | -0.14 | 0.513 |
| Hit rate, delayed recognition | -0.15 | 0.500 | 0.20 | 0.358 | 0.30 | 0.257 | 0.07 | 0.751 | 0.31 | 0.155 | 0.53 | **0.010** | -0.52 | **0.011** |
| False alarm rate, delayed recognition | -0.02 | 0.931 | 0.17 | 0.430 | 0.05 | 0.817 | -0.19 | 0.388 | -0.34 | 0.111 | -0.36 | 0.094 | 0.14 | 0.536 |
| d′, delayed recognition | -0.04 | 0.873 | 0.05 | 0.819 | 0.17 | 0.431 | 0.13 | 0.559 | 0.35 | 0.098 | 0.51 | **0.013** | -0.44 | 0.033 |
| c, delayed recognition | 0.26 | 0.229 | -0.37 | 0.086 | -0.41 | 0.053 | 0.05 | 0.833 | -0.10 | 0.660 | -0.31 | 0.147 | 0.45 | **0.029** |
| RT, immediate face/name | 0.50 | **0.015** | 0.07 | 0.758 | -0.12 | 0.583 | -0.24 | 0.268 | -0.13 | 0.210 | -0.13 | 0.561 | 0.25 | 0.242 |
| RT, delayed face/name | -0.26 | 0.231 | 0.28 | 0.200 | 0.13 | 0.552 | -0.24 | 0.277 | -0.12 | 0.588 | 0.20 | 0.354 | 0.43 | 0.038 |
| RT, immediate face/occupation | -0.18 | 0.424 | 0.44 | 0.035 | 0.19 | 0.380 | -0.42 | 0.046 | -0.49 | **0.017** | -0.23 | 0.285 | 0.29 | 0.172 |
| RT, delayed face/occupation | 0.06 | 0.796 | -0.17 | 0.436 | -0.46 | **0.029** | -0.30 | 0.172 | -0.38 | 0.075 | -0.21 | 0.331 | 0.36 | 0.097 |
| RT, Old items | -0.14 | 0.529 | 0.17 | 0.445 | 0.01 | 0.981 | -0.24 | 0.266 | -0.08 | 0.717 | -0.23 | 0.285 | 0.29 | 0.172 |
| RT, New items | -0.26 | 0.227 | 0.28 | 0.190 | 0.12 | 0.581 | -0.26 | 0.231 | -0.26 | 0.222 | -0.21 | 0.331 | 0.36 | 0.097 |

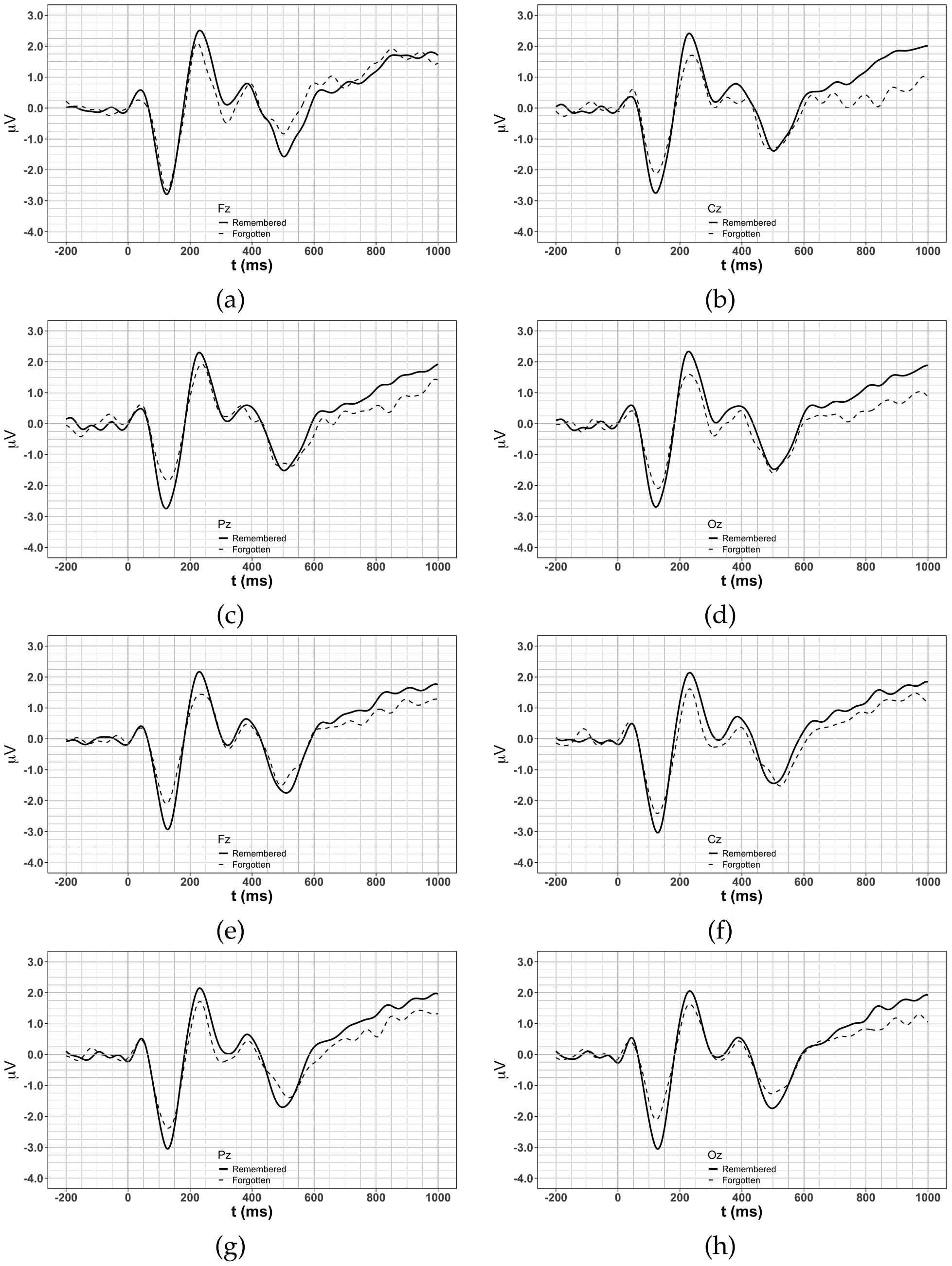

**Figure S4.** Group average wave forms for remembered and forgotten face/name associations: (a) immediate, electrode Fz; (b) immediate, electrode Cz; (c) immediate, electrode Pz; (d) immediate, electrode Oz; (e) delayed, electrode Fz; (f) delayed, electrode Cz; (g) delayed, electrode Pz; (h) delayed, electrode Oz.

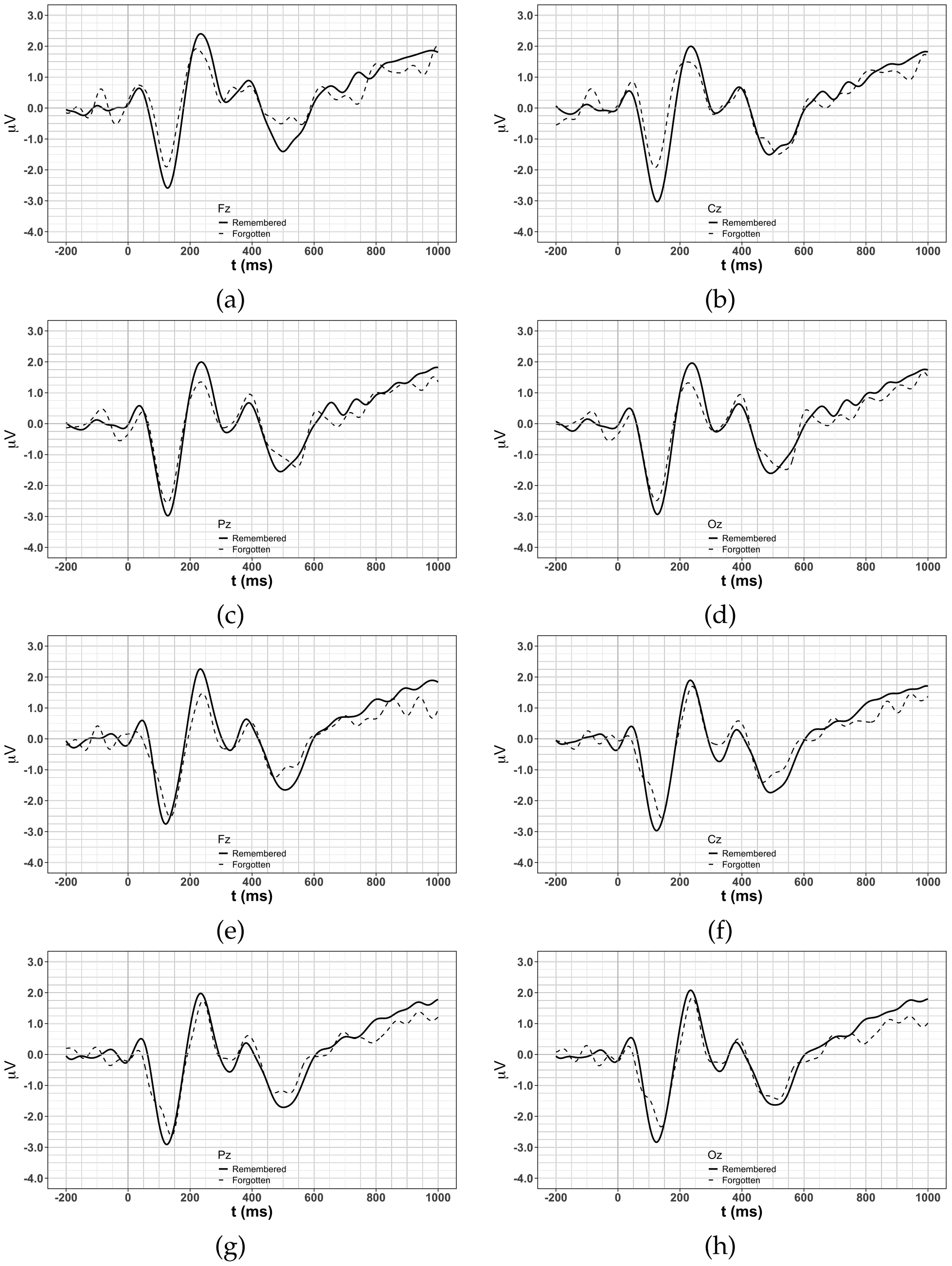

**Figure S5.** Group average wave forms for remembered and forgotten face/occupation associations: (a) immediate, electrode Fz; (b) immediate, electrode Cz; (c) immediate, electrode Pz; (d) immediate, electrode Oz; (e) delayed, electrode Fz; (f) delayed, electrode Cz; (g) delayed, electrode Pz; (h) delayed, electrode Oz.

**Table S6.** Results of the repeated measures analyses of the EEG amplitudes for the FNAM. Cells highlighted in gray indicate statistically significant results.

| Test type | Retention interval | Time interval | Factor | *df* | *F* | *p* |
|---|---|---|---|---|---|---|
| Face/name | Immediate | 200-400 | Electrode (E) | 3 | 1.54 | 0.298 |
| | | | Recall status (R) | 1 | 14.55 | **< 0.001** |
| | | | E x R | 3 | 1.17 | 0.311 |
| | | 400-600 | E | 3 | 4.52 | **0.005** |
| | | | R | 1 | 1.25 | 0.265 |
| | | | E x R | 3 | 0.08 | 0.971 |
| | | 600-800 | E | 3 | 14.55 | **< 0.001** |
| | | | R | 1 | 3.20 | 0.076 |
| | | | E x R | 3 | 0.90 | 0.445 |
| | Delayed | 200-400 | E | 3 | 0.34 | 0.798 |
| | | | R | 1 | 21.09 | **<0.001** |
| | | | E x R | 3 | 0.46 | 0.713 |
| | | 400-600 | E | 3 | 0.10 | 0.960 |
| | | | R | 1 | 0.13 | 0.718 |
| | | | E x R | 3 | 0.62 | 0.606 |
| | | 600-800 | E | 3 | 10.78 | **< 0.001** |
| | | | R | 1 | 0.09 | 0.765 |
| | | | E x R | 3 | 9.14 | **< 0.001** |
| Face/occupation | Immediate | 200-400 | E | 3 | 1.57 | 0.200 |
| | | | R | 1 | 3.41 | 0.067 |
| | | | E x R | 3 | 0.43 | 0.734 |
| | | 400-600 | E | 3 | 0.29 | 0.831 |
| | | | R | 1 | 15.98 | **< 0.001** |
| | | | E x R | 3 | 1.40 | 0.245 |
| | | 600-800 | E | 3 | 5.79 | **0.001** |
| | | | R | 1 | 0.07 | 0.792 |
| | | | E x R | 3 | 5.25 | **0.002** |
| | Delayed | 200-400 | E | 3 | 4.76 | **0.003** |
| | | | R | 1 | 15.76 | **0.001** |
| | | | E x R | 3 | 3.55 | **0.016** |
| | | 400-600 | E | 3 | 8.55 | **0.001** |
| | | | R | 1 | 0.02 | 0.896 |
| | | | E x R | 3 | 6.68 | **0.001** |
| | | 600-800 | E | 3 | 0.56 | 0.645 |
| | | | R | 1 | 2.59 | 0.110 |
| | | | E x R | 3 | 0.02 | 0.996 |

There were three instances in which there was a significant main effect of electrode without there being and electrode × recall status interaction. For main effect of electrode for the face/name associations at the immediate test in the 400-600 ms time interval, the mean (standard error) amplitudes were -0.11 (0.15) $\mu$V at Fz, -0.56 (0.15) $\mu$V at Cz, -0.73 (0.15) $\mu$V at Pz, and -0.71 (0.15) at Oz. For main effect of electrode for the face/name associations at the immediate test in the 600-800 ms time interval, the mean (standard error) amplitudes were 0.89 (0.15) $\mu$V at Fz, 0.42 (0.15) $\mu$V at Cz, 1.18 (0.15) $\mu$V at Pz, and 0.47 (0.15) $\mu$V at Oz. For main effect of electrode for the face/occupation associations at the immediate test in the 600-800 ms time interval, the mean (standard error) amplitudes were 0.64 (0.15) $\mu$V at Fz, 0.45 (0.15) $\mu$V at Cz, 0.97 (0.15) $\mu$V at Pz, and 0.14 (0.15) $\mu$V at Oz.

There were two instances in which there was a main effect for recall status without being involved in the interaction. For the main effect of recall status in face/name recognition at the immediate test in the 200-400 ms time interval, the mean (standard error) amplitudes were 0.83 $\mu$V (0.06) for remembered associations and 0.47 (0.06) $\mu$V for forgotten associations. For the main effect of recall status in face/occupation recognition at the immediate test in the 400-600 ms time interval, the mean (standard error) amplitudes were -1.23 (0.14) $\mu$V for remembered associations and -0.60 (0.14) $\mu$V for forgotten associations.

There were four instances in which there was a electrode × recall status interaction: in face/name recognition at the delayed test in the 600-800 ms time interval, in face/occupation recognition at the immediate test in the 600-800 ms time interval, in face/occupation recognition at the delayed test in the 200-400 ms time interval, and in face/occupation recognition at the delayed test in the 400-600 ms time window. The means for those interactions are plotted in the panels of Supplementary Figure S6.

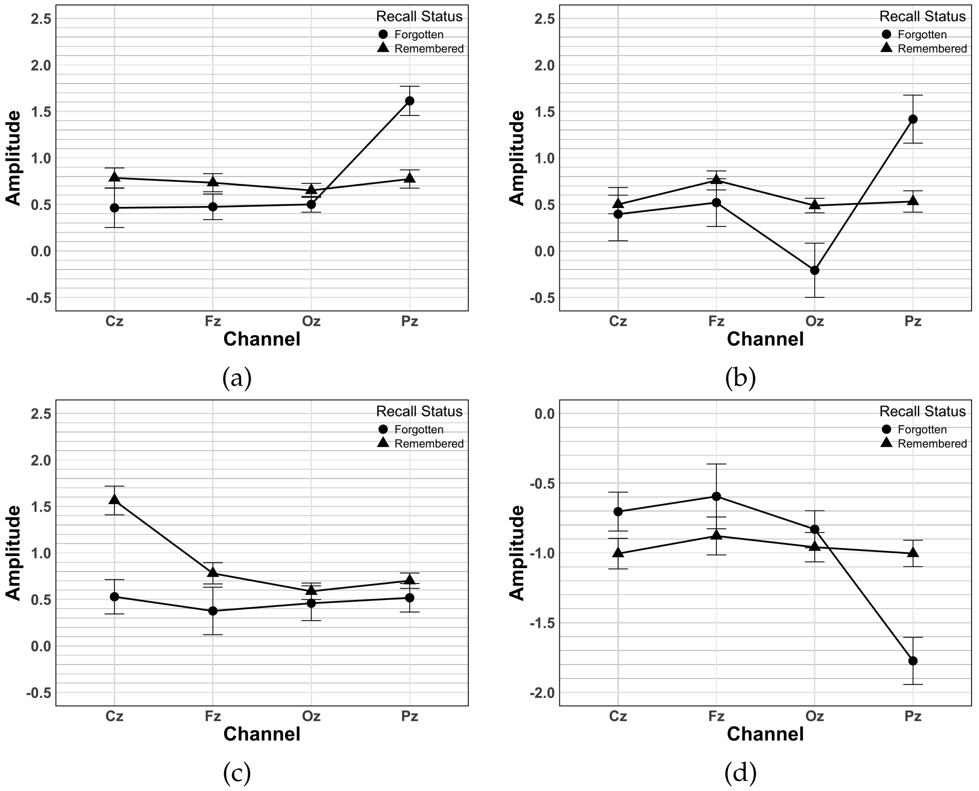

**Figure S6.** Group means for the significant electrode × recall status interactions in the EEG data for the FNAM: (a) face/name delayed 600-800 ms, (b) face/occupation immediate 600-800 ms, (c) face/occupation delayed 200-400 ms, (d) face/occupation delayed 400-600 ms.

#### 4.1.4. Correlations between EEG and iron status

The complete set of correlations between the EEG variables in the conditions of the FNAM and the iron status biomarkers are presented in Supplementary Tables S7-S12.

**Table S7.** Correlations (1) between the EEG amplitudes and iron status biomarkers in the FNAM, for the face/name associations. Cells highlighted in gray indicate correlations that were significant after adjusting for the false discovery rate. Note: pctl = percentile.

| Retention interval | Time interval | Electrode | Recall Status | Age | | Hb | | sFt | | sFt pctl | |
|---|---|---|---|---|---|---|---|---|---|---|---|
| | | | | r | *p* | *r* | *p* | *r* | *p* | *r* | *p* |
| Immediate | 200-400 | Cz | forgotten | 0.30 | 0.211 | -0.18 | 0.456 | -0.05 | 0.844 | -0.16 | 0.506 |
| | | | remembered | 0.13 | 0.592 | 0.29 | 0.224 | 0.69 | **0.001** | 0.50 | **0.029** |
| | | Fz | forgotten | 0.24 | 0.334 | 0.25 | 0.311 | -0.18 | 0.469 | -0.23 | 0.351 |
| | | | remembered | 0.07 | 0.773 | 0.33 | 0.162 | 0.60 | **0.007** | 0.44 | 0.058 |
| | | Oz | forgotten | -0.32 | 0.184 | 0.01 | 0.976 | 0.55 | **0.015** | 0.69 | **0.001** |
| | | | remembered | -0.03 | 0.918 | 0.07 | 0.773 | -0.06 | 0.814 | -0.08 | 0.754 |
| | | Pz | forgotten | 0.28 | 0.253 | -0.14 | 0.563 | -0.07 | 0.768 | -0.19 | 0.428 |
| | | | remembered | 0.06 | 0.795 | -0.12 | 0.640 | -0.18 | 0.474 | -0.23 | 0.354 |
| | 400-600 | Cz | forgotten | -0.10 | 0.678 | -0.15 | 0.536 | -0.15 | 0.544 | -0.09 | 0.726 |
| | | | remembered | 0.22 | 0.363 | 0.18 | 0.472 | -0.62 | **0.005** | -0.69 | **0.001** |
| | | Fz | forgotten | 0.32 | 0.181 | 0.20 | 0.415 | 0.19 | 0.449 | 0.03 | 0.909 |
| | | | remembered | 0.14 | 0.577 | 0.02 | 0.942 | -0.52 | **0.021** | -0.66 | **0.002** |
| | | Oz | forgotten | -0.29 | 0.225 | -0.22 | 0.357 | -0.59 | **0.008** | -0.38 | 0.107 |
| | | | remembered | 0.19 | 0.432 | 0.45 | 0.051 | -0.09 | 0.726 | -0.19 | 0.429 |
| | | Pz | forgotten | -0.15 | 0.554 | -0.24 | 0.318 | -0.09 | 0.719 | -0.01 | 0.960 |
| | | | remembered | 0.26 | 0.282 | 0.20 | 0.419 | -0.08 | 0.750 | -0.19 | 0.429 |
| | 600-800 | Cz | forgotten | -0.17 | 0.498 | -0.26 | 0.288 | -0.08 | 0.751 | -0.05 | 0.844 |
| | | | remembered | -0.19 | 0.433 | 0.37 | 0.125 | 0.44 | 0.058 | 0.47 | 0.044 |
| | | Fz | forgotten | 0.05 | 0.831 | 0.25 | 0.308 | 0.42 | 0.072 | 0.33 | 0.175 |
| | | | remembered | 0.13 | 0.607 | 0.08 | 0.738 | 0.66 | **0.002** | 0.55 | **0.014** |
| | | Oz | forgotten | -0.39 | 0.097 | -0.30 | 0.206 | -0.42 | 0.073 | -0.25 | 0.310 |
| | | | remembered | -0.18 | 0.452 | 0.28 | 0.246 | 0.23 | 0.344 | 0.18 | 0.450 |
| | | Pz | forgotten | -0.17 | 0.500 | 0.47 | 0.042 | 0.19 | 0.447 | 0.23 | 0.345 |
| | | | remembered | -0.23 | 0.352 | 0.25 | 0.293 | 0.29 | 0.237 | 0.26 | 0.277 |
| Delayed | 200-400 | Cz | forgotten | -0.24 | 0.295 | -0.04 | 0.874 | -0.02 | 0.930 | -0.02 | 0.950 |
| | | | remembered | 0.34 | 0.130 | 0.02 | 0.919 | 0.38 | 0.092 | 0.21 | 0.362 |
| | | Fz | forgotten | 0.16 | 0.478 | -0.10 | 0.672 | -0.06 | 0.785 | -0.05 | 0.831 |
| | | | remembered | -0.03 | 0.896 | 0.18 | 0.427 | 0.27 | 0.232 | 0.18 | 0.432 |
| | | Oz | forgotten | 0.11 | 0.626 | 0.01 | 0.955 | 0.70 | **0.000** | 0.67 | **0.001** |
| | | | remembered | 0.05 | 0.824 | -0.27 | 0.244 | 0.51 | **0.017** | 0.42 | 0.056 |
| | | Pz | forgotten | 0.01 | 0.979 | 0.00 | 0.988 | 0.42 | 0.059 | 0.49 | **0.026** |
| | | | remembered | 0.24 | 0.306 | -0.17 | 0.470 | 0.49 | **0.026** | 0.33 | 0.146 |
| | 400-600 | Cz | forgotten | -0.31 | 0.168 | 0.19 | 0.422 | 0.22 | 0.338 | 0.24 | 0.291 |
| | | | remembered | 0.25 | 0.273 | 0.21 | 0.371 | 0.07 | 0.769 | -0.05 | 0.820 |
| | | Fz | forgotten | 0.10 | 0.678 | -0.13 | 0.562 | -0.31 | 0.171 | -0.26 | 0.250 |
| | | | remembered | -0.01 | 0.969 | 0.27 | 0.243 | 0.16 | 0.493 | 0.06 | 0.781 |
| | | Oz | forgotten | -0.39 | 0.084 | -0.08 | 0.718 | -0.11 | 0.643 | 0.03 | 0.904 |
| | | | remembered | 0.09 | 0.706 | -0.13 | 0.566 | -0.78 | **0.000** | -0.74 | **0.000** |
| | | Pz | forgotten | -0.11 | 0.624 | 0.31 | 0.174 | 0.04 | 0.850 | 0.01 | 0.964 |
| | | | remembered | 0.29 | 0.205 | 0.04 | 0.850 | 0.31 | 0.171 | 0.22 | 0.339 |
| | 600-800 | Cz | forgotten | -0.11 | 0.645 | -0.03 | 0.915 | -0.05 | 0.827 | -0.11 | 0.648 |
| | | | remembered | 0.17 | 0.459 | 0.27 | 0.230 | 0.07 | 0.755 | 0.03 | 0.894 |
| | | Fz | forgotten | 0.07 | 0.776 | 0.01 | 0.954 | -0.16 | 0.480 | -0.11 | 0.625 |
| | | | remembered | -0.04 | 0.880 | 0.29 | 0.211 | 0.05 | 0.838 | 0.01 | 0.963 |
| | | Oz | forgotten | -0.42 | 0.059 | -0.28 | 0.226 | -0.25 | 0.273 | -0.08 | 0.735 |
| | | | remembered | -0.09 | 0.695 | 0.36 | 0.111 | 0.29 | 0.203 | 0.35 | 0.125 |
| | | Pz | forgotten | -0.13 | 0.588 | -0.13 | 0.584 | 0.11 | 0.624 | 0.24 | 0.287 |
| | | | remembered | 0.03 | 0.886 | 0.39 | 0.084 | 0.11 | 0.625 | 0.10 | 0.659 |

**Table S8.** Correlations (2) between the EEG amplitudes and iron status biomarkers in the FNAM, for the face/occupation associations. Cells highlighted in gray indicate correlations that were significant after adjusting for the false discovery rate. Note: pctl = percentile

| Retention interval | Time interval | Electrode | Recall Status | Age *r* | Age *p* | Hb *r* | Hb *p* | sFt *r* | sFt *p* | sFt pctl *r* | sFt pctl *p* |
|---|---|---|---|---|---|---|---|---|---|---|---|
| Immediate | 209-400 | Cz | forgotten | 0.06 | 0.809 | 0.27 | 0.273 | -0.07 | 0.770 | -0.16 | 0.509 |
| | | | remembered | 0.36 | 0.134 | -0.10 | 0.679 | -0.32 | 0.179 | -0.46 | 0.050 |
| | | Fz | forgotten | 0.28 | 0.252 | 0.16 | 0.526 | -0.29 | 0.237 | -0.40 | 0.088 |
| | | | remembered | 0.01 | 0.963 | 0.26 | 0.292 | 0.24 | 0.323 | 0.26 | 0.280 |
| | | Oz | forgotten | -0.04 | 0.888 | 0.17 | 0.489 | -0.37 | 0.121 | -0.34 | 0.161 |
| | | | remembered | -0.41 | 0.082 | 0.15 | 0.529 | 0.21 | 0.396 | 0.35 | 0.141 |
| | | Pz | forgotten | -0.02 | 0.930 | 0.21 | 0.387 | -0.27 | 0.258 | -0.27 | 0.263 |
| | | | remembered | -0.43 | 0.068 | 0.25 | 0.311 | 0.14 | 0.569 | 0.31 | 0.205 |
| | 400-600 | Cz | forgotten | 0.07 | 0.772 | 0.33 | 0.162 | 0.01 | 0.956 | -0.03 | 0.892 |
| | | | remembered | 0.09 | 0.708 | -0.22 | 0.372 | -0.10 | 0.698 | -0.14 | 0.571 |
| | | Fz | forgotten | 0.34 | 0.159 | 0.45 | 0.054 | -0.01 | 0.984 | -0.14 | 0.566 |
| | | | remembered | -0.03 | 0.892 | -0.31 | 0.200 | -0.04 | 0.887 | -0.01 | 0.975 |
| | | Oz | forgotten | -0.09 | 0.726 | 0.36 | 0.135 | -0.37 | 0.124 | -0.30 | 0.215 |
| | | | remembered | 0.19 | 0.438 | -0.35 | 0.139 | 0.04 | 0.881 | -0.02 | 0.936 |
| | | Pz | forgotten | -0.06 | 0.794 | 0.40 | 0.089 | -0.25 | 0.307 | -0.22 | 0.365 |
| | | | remembered | 0.15 | 0.535 | -0.37 | 0.117 | 0.10 | 0.694 | 0.02 | 0.944 |
| | 600-800 | Cz | forgotten | 0.04 | 0.860 | 0.14 | 0.576 | -0.26 | 0.287 | -0.32 | 0.185 |
| | | | remembered | -0.30 | 0.210 | -0.20 | 0.412 | 0.25 | 0.301 | 0.36 | 0.127 |
| | | Fz | forgotten | 0.15 | 0.548 | 0.02 | 0.946 | 0.44 | 0.058 | 0.53 | **0.019** |
| | | | remembered | 0.06 | 0.822 | 0.32 | 0.184 | -0.10 | 0.689 | -0.08 | 0.757 |
| | | Oz | forgotten | -0.10 | 0.686 | 0.45 | 0.053 | 0.77 | **0.000** | 0.76 | **0.000** |
| | | | remembered | -0.21 | 0.400 | 0.01 | 0.956 | 0.16 | 0.521 | 0.29 | 0.236 |
| | | Pz | forgotten | 0.14 | 0.558 | 0.10 | 0.687 | 0.66 | **0.002** | 0.60 | **0.006** |
| | | | remembered | -0.13 | 0.589 | 0.14 | 0.575 | 0.20 | 0.403 | 0.31 | 0.198 |
| Delayed | 200-400 | Cz | forgotten | 0.11 | 0.640 | -0.13 | 0.568 | 0.27 | 0.243 | 0.22 | 0.340 |
| | | | remembered | 0.09 | 0.692 | 0.48 | **0.027** | -0.28 | 0.226 | -0.29 | 0.208 |
| | | Fz | forgotten | 0.01 | 0.983 | 0.02 | 0.935 | 0.09 | 0.687 | 0.10 | 0.662 |
| | | | remembered | 0.25 | 0.266 | -0.01 | 0.953 | 0.11 | 0.644 | 0.00 | 0.995 |
| | | Oz | forgotten | 0.02 | 0.934 | 0.16 | 0.490 | -0.13 | 0.589 | -0.10 | 0.679 |
| | | | remembered | 0.10 | 0.683 | -0.16 | 0.500 | 0.03 | 0.908 | -0.08 | 0.724 |
| | | Pz | forgotten | 0.09 | 0.690 | -0.01 | 0.971 | 0.23 | 0.313 | 0.19 | 0.398 |
| | | | remembered | 0.12 | 0.606 | 0.40 | 0.069 | -0.07 | 0.753 | -0.04 | 0.855 |
| | 400-600 | Cz | forgotten | 0.02 | 0.917 | -0.28 | 0.222 | -0.49 | **0.026** | 0.41 | 0.069 |
| | | | remembered | 0.33 | 0.149 | -0.53 | **0.014** | 0.19 | 0.401 | 0.10 | 0.667 |
| | | Fz | forgotten | -0.09 | 0.701 | 0.05 | 0.832 | 0.29 | 0.199 | 0.31 | 0.178 |
| | | | remembered | 0.48 | **0.026** | 0.19 | 0.403 | -0.04 | 0.857 | -0.22 | 0.345 |
| | | Oz | forgotten | 0.02 | 0.949 | -0.07 | 0.749 | 0.32 | 0.160 | 0.24 | 0.290 |
| | | | remembered | 0.27 | 0.230 | -0.22 | 0.342 | -0.01 | 0.954 | -0.14 | 0.555 |
| | | Pz | forgotten | -0.04 | 0.861 | 0.18 | 0.438 | -0.63 | **0.003** | -0.55 | **0.012** |
| | | | remembered | 0.28 | 0.223 | -0.38 | 0.088 | 0.05 | 0.836 | -0.05 | 0.835 |
| | 600-800 | Cz | forgotten | -0.11 | 0.632 | -0.23 | 0.314 | 0.09 | 0.706 | 0.10 | 0.659 |
| | | | remembered | 0.34 | 0.129 | -0.08 | 0.726 | 0.04 | 0.860 | -0.03 | 0.904 |
| | | Fz | forgotten | -0.12 | 0.619 | 0.10 | 0.675 | -0.11 | 0.628 | -0.05 | 0.848 |
| | | | remembered | 0.33 | 0.139 | 0.47 | 0.032 | -0.15 | 0.521 | -0.28 | 0.218 |
| | | Oz | forgotten | -0.20 | 0.390 | 0.08 | 0.741 | -0.26 | 0.256 | -0.19 | 0.408 |
| | | | remembered | 0.13 | 0.567 | 0.05 | 0.825 | -0.10 | 0.674 | -0.21 | 0.352 |
| | | Pz | forgotten | -0.12 | 0.602 | -0.07 | 0.771 | -0.08 | 0.720 | -0.05 | 0.837 |
| | | | remembered | 0.07 | 0.755 | 0.00 | 0.988 | -0.03 | 0.898 | -0.08 | 0.724 |

**Table S9.** Correlations (3) between the EEG amplitudes and iron status biomarkers in the FNAM, for the face/name associations. Cells highlighted in gray indicate correlations that were significant after adjusting for the false discovery rate.

| Retention interval | Time Bin | Electrode | Recall Status | RBC | | HCT | | RDW | |
|---|---|---|---|---|---|---|---|---|---|
| | | | | *r* | *p* | *r* | *p* | *r* | *p* |
| Immediate | 200-400 | Cz | forgotten | -0.08 | 0.757 | -0.09 | 0.714 | 0.27 | 0.256 |
| | | | remembered | 0.23 | 0.342 | 0.30 | 0.218 | -0.15 | 0.543 |
| | | Fz | forgotten | 0.22 | 0.356 | 0.20 | 0.422 | -0.17 | 0.493 |
| | | | remembered | 0.25 | 0.310 | 0.31 | 0.201 | -0.18 | 0.471 |
| | | Oz | forgotten | 0.01 | 0.959 | -0.02 | 0.924 | -0.41 | 0.078 |
| | | | remembered | 0.22 | 0.371 | 0.12 | 0.627 | 0.24 | 0.318 |
| | | Pz | forgotten | -0.10 | 0.689 | -0.09 | 0.705 | 0.28 | 0.249 |
| | | | remembered | -0.08 | 0.732 | -0.07 | 0.775 | 0.36 | 0.133 |
| | 400-600 | Cz | forgotten | -0.25 | 0.311 | -0.16 | 0.505 | 0.10 | 0.685 |
| | | | remembered | 0.09 | 0.718 | 0.05 | 0.853 | 0.36 | 0.136 |
| | | Fz | forgotten | 0.23 | 0.342 | 0.29 | 0.222 | -0.05 | 0.849 |
| | | | remembered | -0.05 | 0.836 | -0.04 | 0.885 | 0.45 | 0.053 |
| | | Oz | forgotten | -0.36 | 0.132 | -0.33 | 0.162 | 0.01 | 0.963 |
| | | | remembered | 0.29 | 0.223 | 0.47 | 0.041 | -0.11 | 0.649 |
| | | Pz | forgotten | -0.34 | 0.153 | -0.26 | 0.287 | 0.04 | 0.864 |
| | | | remembered | 0.02 | 0.953 | 0.17 | 0.490 | -0.06 | 0.804 |
| | 600-800 | Cz | forgotten | -0.35 | 0.148 | -0.23 | 0.341 | 0.20 | 0.415 |
| | | | remembered | 0.26 | 0.278 | 0.28 | 0.245 | -0.20 | 0.421 |
| | | Fz | forgotten | 0.31 | 0.204 | 0.37 | 0.125 | -0.12 | 0.633 |
| | | | remembered | 0.11 | 0.652 | 0.14 | 0.577 | -0.27 | 0.258 |
| | | Oz | forgotten | -0.47 | 0.041 | -0.40 | 0.090 | 0.06 | 0.824 |
| | | | remembered | 0.23 | 0.352 | 0.32 | 0.179 | -0.08 | 0.738 |
| | | Pz | forgotten | 0.48 | 0.038 | 0.42 | 0.074 | -0.14 | 0.573 |
| | | | remembered | 0.16 | 0.514 | 0.24 | 0.314 | -0.13 | 0.610 |
| Delayed | 200-400 | Cz | forgotten | 0.03 | 0.897 | 0.02 | 0.922 | 0.09 | 0.715 |
| | | | remembered | -0.02 | 0.920 | 0.08 | 0.730 | -0.08 | 0.748 |
| | | Fz | forgotten | -0.22 | 0.347 | -0.15 | 0.518 | 0.09 | 0.710 |
| | | | remembered | 0.05 | 0.818 | 0.19 | 0.413 | -0.06 | 0.814 |
| | | Oz | forgotten | 0.10 | 0.676 | -0.05 | 0.841 | -0.19 | 0.413 |
| | | | remembered | -0.32 | 0.153 | -0.19 | 0.408 | -0.12 | 0.617 |
| | | Pz | forgotten | 0.06 | 0.786 | 0.05 | 0.839 | 0.08 | 0.746 |
| | | | remembered | -0.18 | 0.430 | -0.07 | 0.768 | -0.03 | 0.892 |
| | 400-600 | Cz | forgotten | 0.22 | 0.346 | 0.26 | 0.255 | -0.08 | 0.741 |
| | | | remembered | 0.06 | 0.792 | 0.20 | 0.389 | -0.02 | 0.936 |
| | | Fz | forgotten | -0.22 | 0.334 | -0.19 | 0.415 | 0.21 | 0.372 |
| | | | remembered | 0.04 | 0.870 | 0.23 | 0.314 | -0.05 | 0.823 |
| | | Oz | forgotten | -0.26 | 0.260 | -0.19 | 0.406 | 0.05 | 0.819 |
| | | | remembered | -0.19 | 0.409 | -0.19 | 0.408 | 0.29 | 0.201 |
| | | Pz | forgotten | 0.34 | 0.127 | 0.37 | 0.099 | -0.14 | 0.534 |
| | | | remembered | 0.04 | 0.873 | 0.07 | 0.760 | -0.11 | 0.641 |
| | 600-800 | Cz | forgotten | 0.01 | 0.966 | 0.08 | 0.728 | 0.15 | 0.523 |
| | | | remembered | 0.26 | 0.254 | 0.36 | 0.106 | -0.15 | 0.504 |
| | | Fz | forgotten | -0.11 | 0.624 | -0.07 | 0.753 | 0.03 | 0.913 |
| | | | remembered | 0.17 | 0.461 | 0.31 | 0.169 | -0.14 | 0.541 |
| | | Oz | forgotten | -0.34 | 0.129 | -0.30 | 0.192 | 0.09 | 0.711 |
| | | | remembered | 0.39 | 0.078 | 0.37 | 0.102 | -0.27 | 0.236 |
| | | Pz | forgotten | -0.11 | 0.628 | -0.17 | 0.458 | -0.03 | 0.910 |
| | | | remembered | 0.43 | 0.054 | 0.44 | 0.045 | -0.11 | 0.644 |

**Table S10.** Correlations (4) between the EEG amplitudes and iron status biomarkers in the FNAM, for the face/occupation associations. Cells highlighted in gray indicate correlations that were significant after adjusting for the false discovery rate.

| **Retention interval** | **Time Bin** | **Electrode** | **Recall Status** | **RBC** | | **HCT** | | **RDW** | |
|---|---|---|---|---|---|---|---|---|---|
| | | | | *r* | *p* | *r* | *p* | *r* | *p* |
| Immediate | 200-400 | Cz | forgotten | 0.14 | 0.556 | 0.30 | 0.206 | 0.17 | 0.498 |
| | | | remembered | -0.15 | 0.544 | -0.01 | 0.956 | 0.26 | 0.290 |
| | | Fz | forgotten | 0.14 | 0.567 | 0.21 | 0.394 | 0.12 | 0.624 |
| | | | remembered | 0.23 | 0.345 | 0.09 | 0.712 | -0.29 | 0.236 |
| | | Oz | forgotten | 0.05 | 0.848 | 0.17 | 0.485 | 0.16 | 0.507 |
| | | | remembered | 0.27 | 0.273 | 0.08 | 0.734 | -0.21 | 0.382 |
| | | Pz | forgotten | 0.12 | 0.621 | 0.24 | 0.320 | 0.24 | 0.323 |
| | | | remembered | 0.31 | 0.201 | 0.18 | 0.470 | -0.19 | 0.425 |
| | 400-600 | Cz | forgotten | 0.22 | 0.375 | 0.29 | 0.236 | -0.02 | 0.929 |
| | | | remembered | -0.21 | 0.378 | -0.05 | 0.855 | -0.01 | 0.970 |
| | | Fz | forgotten | 0.35 | 0.141 | 0.46 | 0.049 | -0.19 | 0.449 |
| | | | remembered | -0.30 | 0.220 | -0.12 | 0.624 | 0.02 | 0.947 |
| | | Oz | forgotten | 0.19 | 0.442 | 0.27 | 0.256 | 0.06 | 0.800 |
| | | | remembered | -0.41 | 0.080 | -0.21 | 0.398 | 0.03 | 0.903 |
| | | Pz | forgotten | 0.26 | 0.290 | 0.34 | 0.152 | 0.13 | 0.608 |
| | | | remembered | -0.43 | 0.069 | -0.18 | 0.451 | -0.13 | 0.590 |
| | 600-800 | Cz | forgotten | -0.03 | 0.910 | 0.09 | 0.712 | 0.22 | 0.378 |
| | | | remembered | -0.31 | 0.193 | -0.26 | 0.281 | -0.02 | 0.929 |
| | | Fz | forgotten | -0.05 | 0.844 | 0.03 | 0.920 | 0.20 | 0.406 |
| | | | remembered | 0.12 | 0.622 | 0.09 | 0.707 | -0.22 | 0.360 |
| | | Oz | forgotten | 0.33 | 0.163 | 0.46 | 0.050 | -0.45 | 0.054 |
| | | | remembered | -0.06 | 0.825 | -0.17 | 0.478 | -0.09 | 0.712 |
| | | Pz | forgotten | 0.13 | 0.601 | 0.18 | 0.459 | -0.42 | 0.073 |
| | | | remembered | 0.07 | 0.791 | -0.03 | 0.893 | 0.02 | 0.937 |
| Delayed | 200-400 | Cz | forgotten | 0.01 | 0.958 | -0.06 | 0.794 | 0.09 | 0.690 |
| | | | remembered | 0.38 | 0.087 | 0.39 | 0.083 | -0.04 | 0.860 |
| | | Fz | forgotten | 0.17 | 0.453 | 0.16 | 0.495 | 0.04 | 0.882 |
| | | | remembered | -0.17 | 0.452 | 0.07 | 0.749 | -0.22 | 0.336 |
| | | Oz | forgotten | 0.23 | 0.308 | 0.14 | 0.554 | 0.07 | 0.753 |
| | | | remembered | -0.25 | 0.272 | -0.13 | 0.567 | -0.05 | 0.841 |
| | | Pz | forgotten | 0.08 | 0.725 | 0.02 | 0.947 | 0.12 | 0.599 |
| | | | remembered | 0.44 | 0.048 | 0.33 | 0.149 | -0.10 | 0.655 |
| | 400-600 | Cz | forgotten | -0.18 | 0.425 | -0.11 | 0.652 | 0.02 | 0.940 |
| | | | remembered | -0.41 | 0.065 | -0.42 | 0.055 | 0.00 | 0.994 |
| | | Fz | forgotten | -0.04 | 0.869 | 0.19 | 0.401 | -0.35 | 0.118 |
| | | | remembered | 0.01 | 0.951 | 0.22 | 0.346 | -0.14 | 0.559 |
| | | Oz | forgotten | 0.05 | 0.818 | 0.07 | 0.753 | 0.12 | 0.602 |
| | | | remembered | -0.17 | 0.473 | -0.18 | 0.448 | 0.09 | 0.700 |
| | | Pz | forgotten | 0.07 | 0.784 | 0.01 | 0.980 | 0.01 | 0.955 |
| | | | remembered | -0.31 | 0.177 | -0.35 | 0.120 | 0.10 | 0.676 |
| | 600-800 | Cz | forgotten | -0.07 | 0.754 | -0.08 | 0.741 | 0.16 | 0.481 |
| | | | remembered | -0.14 | 0.552 | -0.12 | 0.600 | -0.25 | 0.274 |
| | | Fz | forgotten | 0.09 | 0.702 | 0.22 | 0.350 | -0.21 | 0.371 |
| | | | remembered | 0.16 | 0.496 | 0.43 | 0.050 | -0.21 | 0.356 |
| | | Oz | forgotten | 0.12 | 0.592 | 0.15 | 0.510 | 0.13 | 0.574 |
| | | | remembered | -0.16 | 0.497 | -0.03 | 0.886 | -0.05 | 0.847 |
| | | Pz | forgotten | 0.07 | 0.772 | 0.07 | 0.758 | 0.18 | 0.444 |
| | | | remembered | -0.22 | 0.349 | -0.16 | 0.492 | -0.11 | 0.633 |

**Table S11.** Correlations (5) between the EEG amplitudes and iron status biomarkers in the FNAM, for the face/name associations. Cells highlighted in gray indicate correlations that were significant after adjusting for the false discovery rate.

| Retention interval | Time Bin | Electrode | Recall Status | MCV | | MCH | | MCHC | |
|---|---|---|---|---|---|---|---|---|---|
| | | | | *r* | *p* | *r* | *p* | *r* | *p* |
| Immediate | 200-400 | Cz | forgotten | 0.00 | 0.996 | -0.14 | 0.565 | -0.28 | 0.242 |
| | | | remembered | 0.03 | 0.903 | 0.08 | 0.736 | 0.11 | 0.643 |
| | | Fz | forgotten | -0.11 | 0.653 | -0.01 | 0.960 | 0.19 | 0.442 |
| | | | remembered | 0.01 | 0.961 | 0.11 | 0.661 | 0.20 | 0.410 |
| | | Oz | forgotten | -0.08 | 0.744 | -0.04 | 0.888 | 0.09 | 0.725 |
| | | | remembered | -0.17 | 0.492 | -0.20 | 0.414 | -0.09 | 0.731 |
| | | Pz | forgotten | 0.03 | 0.9 | -0.05 | 0.833 | -0.16 | 0.503 |
| | | | remembered | 0.06 | 0.823 | -0.03 | 0.917 | -0.14 | 0.557 |
| | 400-600 | Cz | forgotten | 0.17 | 0.476 | 0.14 | 0.555 | -0.02 | 0.941 |
| | | | remembered | -0.08 | 0.75 | 0.10 | 0.686 | 0.35 | 0.137 |
| | | Fz | forgotten | -0.01 | 0.982 | -0.09 | 0.720 | -0.16 | 0.512 |
| | | | remembered | 0.05 | 0.838 | 0.10 | 0.688 | 0.11 | 0.648 |
| | | Oz | forgotten | 0.12 | 0.623 | 0.20 | 0.417 | 0.18 | 0.461 |
| | | | remembered | 0.15 | 0.553 | 0.19 | 0.449 | 0.11 | 0.643 |
| | | Pz | forgotten | 0.21 | 0.384 | 0.17 | 0.479 | -0.04 | 0.863 |
| | | | remembered | 0.19 | 0.448 | 0.23 | 0.341 | 0.14 | 0.581 |
| | 600-800 | Cz | forgotten | 0.25 | 0.303 | 0.15 | 0.534 | -0.16 | 0.520 |
| | | | remembered | -0.06 | 0.822 | 0.12 | 0.641 | 0.34 | 0.154 |
| | | Fz | forgotten | -0.02 | 0.946 | -0.11 | 0.643 | -0.19 | 0.435 |
| | | | remembered | -0.02 | 0.952 | -0.06 | 0.811 | -0.09 | 0.720 |
| | | Oz | forgotten | 0.24 | 0.329 | 0.28 | 0.247 | 0.11 | 0.654 |
| | | | remembered | 0.06 | 0.812 | 0.05 | 0.831 | 0.01 | 0.975 |
| | | Pz | forgotten | -0.22 | 0.363 | -0.06 | 0.795 | 0.29 | 0.229 |
| | | | remembered | 0.07 | 0.792 | 0.11 | 0.642 | 0.12 | 0.623 |
| Delayed | 200-400 | Cz | forgotten | -0.02 | 0.924 | -0.10 | 0.680 | -0.17 | 0.456 |
| | | | remembered | 0.16 | 0.478 | 0.09 | 0.697 | -0.12 | 0.597 |
| | | Fz | forgotten | 0.14 | 0.556 | 0.17 | 0.451 | 0.11 | 0.641 |
| | | | remembered | 0.17 | 0.466 | 0.18 | 0.445 | 0.05 | 0.826 |
| | | Oz | forgotten | -0.25 | 0.27 | -0.15 | 0.513 | 0.18 | 0.448 |
| | | | remembered | 0.27 | 0.241 | 0.12 | 0.617 | -0.27 | 0.233 |
| | | Pz | forgotten | -0.03 | 0.903 | -0.08 | 0.733 | -0.13 | 0.591 |
| | | | remembered | 0.22 | 0.348 | 0.06 | 0.810 | -0.30 | 0.193 |
| | 400-600 | Cz | forgotten | 0.00 | 0.995 | -0.06 | 0.814 | -0.12 | 0.609 |
| | | | remembered | 0.15 | 0.522 | 0.17 | 0.460 | 0.09 | 0.710 |
| | | Fz | forgotten | 0.09 | 0.705 | 0.12 | 0.615 | 0.08 | 0.724 |
| | | | remembered | 0.23 | 0.313 | 0.29 | 0.198 | 0.18 | 0.440 |
| | | Oz | forgotten | 0.15 | 0.523 | 0.23 | 0.309 | 0.21 | 0.366 |
| | | | remembered | 0.05 | 0.837 | 0.08 | 0.729 | 0.06 | 0.792 |
| | | Pz | forgotten | -0.04 | 0.853 | -0.06 | 0.801 | -0.04 | 0.853 |
| | | | remembered | 0.03 | 0.916 | -0.01 | 0.972 | -0.05 | 0.817 |
| | 600-800 | Cz | forgotten | 0.09 | 0.708 | -0.05 | 0.818 | -0.27 | 0.231 |
| | | | remembered | 0.07 | 0.769 | 0.02 | 0.944 | -0.10 | 0.677 |
| | | Fz | forgotten | 0.08 | 0.737 | 0.18 | 0.426 | 0.23 | 0.319 |
| | | | remembered | 0.14 | 0.533 | 0.16 | 0.489 | 0.05 | 0.818 |
| | | Oz | forgotten | 0.15 | 0.53 | 0.10 | 0.679 | -0.08 | 0.728 |
| | | | remembered | -0.14 | 0.544 | -0.06 | 0.788 | 0.13 | 0.578 |
| | | Pz | forgotten | -0.05 | 0.833 | -0.01 | 0.968 | 0.07 | 0.761 |
| | | | remembered | -0.09 | 0.705 | -0.07 | 0.759 | 0.01 | 0.953 |

**Table S12.** Correlations (6) between the EEG amplitudes and iron status biomarkers in the FNAM, for the face/occupation associations. Cells highlighted in gray indicate correlations that were significant after adjusting for the false discovery rate.

| **Retention interval** | **Time Bin** | **Electrode** | **Recall Status** | **MCV** | | **MCH** | | **MCHC** | |
|---|---|---|---|---|---|---|---|---|---|
| | | | | *r* | *p* | *r* | *p* | *r* | *p* |
| Immediate | 200-400 | Cz | forgotten | 0.17 | 0.476 | 0.16 | 0.528 | -0.01 | 0.976 |
| | | | remembered | 0.20 | 0.404 | 0.06 | 0.795 | -0.25 | 0.302 |
| | | Fz | forgotten | 0.05 | 0.846 | 0.00 | 0.996 | -0.08 | 0.738 |
| | | | remembered | -0.25 | 0.313 | 0.02 | 0.943 | 0.49 | 0.034 |
| | | Oz | forgotten | 0.16 | 0.505 | 0.18 | 0.468 | 0.05 | 0.842 |
| | | | remembered | -0.31 | 0.195 | -0.17 | 0.479 | 0.23 | 0.344 |
| | | Pz | forgotten | 0.14 | 0.565 | 0.13 | 0.602 | -0.01 | 0.977 |
| | | | remembered | -0.26 | 0.287 | -0.11 | 0.655 | 0.26 | 0.282 |
| | 400-600 | Cz | forgotten | 0.03 | 0.889 | 0.14 | 0.564 | 0.22 | 0.360 |
| | | | remembered | 0.28 | 0.253 | 0.02 | 0.925 | -0.48 | 0.039 |
| | | Fz | forgotten | 0.02 | 0.922 | 0.08 | 0.736 | 0.13 | 0.596 |
| | | | remembered | 0.32 | 0.188 | 0.02 | 0.923 | -0.54 | **0.016** |
| | | Oz | forgotten | 0.07 | 0.78 | 0.22 | 0.366 | 0.31 | 0.194 |
| | | | remembered | 0.39 | 0.096 | 0.14 | 0.583 | -0.48 | 0.039 |
| | | Pz | forgotten | 0.05 | 0.843 | 0.18 | 0.461 | 0.27 | 0.260 |
| | | | remembered | 0.44 | 0.062 | 0.12 | 0.612 | -0.57 | **0.011** |
| | 600-800 | Cz | forgotten | 0.16 | 0.504 | 0.22 | 0.362 | 0.15 | 0.554 |
| | | | remembered | 0.19 | 0.443 | 0.22 | 0.375 | 0.08 | 0.747 |
| | | Fz | forgotten | 0.11 | 0.653 | 0.09 | 0.723 | -0.04 | 0.884 |
| | | | remembered | -0.08 | 0.735 | 0.25 | 0.303 | 0.65 | **0.003** |
| | | Oz | forgotten | 0.04 | 0.878 | 0.12 | 0.635 | 0.17 | 0.484 |
| | | | remembered | -0.14 | 0.579 | 0.10 | 0.686 | 0.46 | 0.050 |
| | | Pz | forgotten | 0.01 | 0.957 | -0.06 | 0.810 | -0.13 | 0.584 |
| | | | remembered | -0.15 | 0.552 | 0.10 | 0.695 | 0.46 | 0.049 |
| Delayed | 200-400 | Cz | forgotten | -0.06 | 0.793 | -0.15 | 0.505 | -0.20 | 0.394 |
| | | | remembered | -0.12 | 0.594 | 0.08 | 0.746 | 0.40 | 0.075 |
| | | Fz | forgotten | -0.04 | 0.848 | -0.19 | 0.405 | -0.32 | 0.155 |
| | | | remembered | 0.38 | 0.094 | 0.24 | 0.303 | -0.21 | 0.354 |
| | | Oz | forgotten | -0.16 | 0.481 | -0.09 | 0.701 | 0.11 | 0.639 |
| | | | remembered | 0.21 | 0.357 | 0.15 | 0.529 | -0.11 | 0.637 |
| | | Pz | forgotten | -0.07 | 0.76 | -0.09 | 0.706 | -0.04 | 0.849 |
| | | | remembered | -0.27 | 0.23 | -0.09 | 0.688 | 0.34 | 0.137 |
| | 400-600 | Cz | forgotten | 0.19 | 0.408 | -0.08 | 0.740 | -0.50 | **0.021** |
| | | | remembered | 0.11 | 0.649 | -0.12 | 0.609 | -0.45 | 0.043 |
| | | Fz | forgotten | 0.33 | 0.147 | 0.15 | 0.525 | -0.33 | 0.150 |
| | | | remembered | 0.25 | 0.269 | 0.23 | 0.319 | 0.01 | 0.982 |
| | | Oz | forgotten | 0.04 | 0.861 | -0.15 | 0.522 | -0.38 | 0.092 |
| | | | remembered | 0.02 | 0.944 | -0.08 | 0.730 | -0.20 | 0.393 |
| | | Pz | forgotten | -0.11 | 0.639 | 0.14 | 0.566 | 0.47 | 0.036 |
| | | | remembered | 0.03 | 0.914 | -0.09 | 0.698 | -0.24 | 0.304 |
| | 600-800 | Cz | forgotten | 0.05 | 0.82 | -0.16 | 0.484 | -0.44 | 0.046 |
| | | | remembered | 0.08 | 0.737 | 0.11 | 0.640 | 0.06 | 0.790 |
| | | Fz | forgotten | 0.16 | 0.483 | 0.04 | 0.874 | -0.25 | 0.279 |
| | | | remembered | 0.31 | 0.169 | 0.41 | 0.063 | 0.26 | 0.259 |
| | | Oz | forgotten | 0.03 | 0.913 | -0.04 | 0.852 | -0.16 | 0.496 |
| | | | remembered | 0.21 | 0.362 | 0.31 | 0.179 | 0.23 | 0.327 |
| | | Pz | forgotten | 0.02 | 0.935 | -0.15 | 0.522 | -0.36 | 0.114 |
| | | | remembered | 0.16 | 0.483 | 0.34 | 0.127 | 0.38 | 0.086 |

### *4.2. PST*

#### 4.2.1. Behavioral data

Descriptive statistics for the behavioral data from the PST are presented in Supplementary Table S13. Accuracy data (choosing A vs. avoiding B) were analyzed using a one-way repeated measures mixed models ANOVA, with choice type (choose A, avoid B) manipulated within participants and participants as the random factor, and the results are presented in Supplementary Table S14. There was no significant effect of choice type on accuracy, indicating that participants were equally accurate in correctly choosing A (M = .78, SE = .03) vs. avoiding B (M = .79, SE = .03). This aligns with previous literature, suggesting that healthy individuals do not show a difference in either approach or avoidance learning [5,6].

**Table S13.** Descriptive statistics for the behavioral data from the PST.

| Factor | DV | Level | *M* | *SE* | *min* | *max* |
|---|---|---|---|---|---|---|
| Choice type | P(C) | Choose A | 0.78 | 0.03 | 0.36 | 1 |
| | | Avoid B | 0.79 | 0.03 | 0.52 | 1 |
| Conflict Level | RT (ms) | Low | 1027 | 50 | 647 | 1521 |
| | | High | 1311 | 67 | 752 | 1962 |

**Table S14.** Results of the repeated measures ANOVA on the behavioral variables from the PST. Cells highlighted in gray indicate statistically significant effects.

| Factor | DV | *F* | *p* |
|---|---|---|---|
| Choice Type | Accuracy, P(C) | 0.16 | 0.691 |
| Conflict Level | RT, ms | 23.07 | < 0.001 |

A one-way repeated measures ANOVA was performed on correct RTs with conflict level (low, high) manipulated within participants and participants as the random factor. This analysis (see Supplementary Table S14) revealed a main effect for level of conflict, with participants responding more quickly in the low conflict condition (M = 1027, SE = 50) relative to the high conflict condition (M = 1311, SE = 67). These results agree with previous literature, suggesting a conflict-induced slowing in healthy adults [6].

Pearson correlations were calculated involving response accuracy for the two choices and the blood iron biomarkers, and the results are presented in Tables S15 and **??**. Levels of sFt were positively correlated with accuracy of choosing A. Accuracy of correctly avoiding B was positively correlated with sFt and sFt percentile. The results with sFt are consistent with the idea that higher levels of iron should be associated with better dopaminergic system integrity, with the latter being associated with higher levels of performance [5,21].

Pearson correlations were calculated for correct RTs for the two conflict levels (low vs. high) and the iron biomarkers and age (Supplementary Tables S15 and **??**). RTs in the high conflict condition were negatively correlated with sFt, sFt percentile and with MCH. According to Frank et al. [6], participants with higher DA levels showed shorter RTs. Given the evidence in animal models of relationship between iron levels and central, this suggests that even in women who are not iron deficient there still may be a positive relationship between iron status and DA. In addition the relationship with MCH again points to the potential involvement of oxygen transport in non-anemic women.

**Table S15.** Correlations of the behavioral variables in the PST with the iron status biomarkers (1). Cells highlighted in gray indicate correlations that were significant after correcting for the false discovery rate. Note: pctl = percentile.

| | Hb | | sFt | | sFt pctl | | Age | |
|---|---|---|---|---|---|---|---|---|
| Variable | *r* | *p* | *r* | *p* | *r* | *p* | *r* | *p* |
| Choose A, P(C) | 0.40 | 0.047 | 0.46 | **0.022** | 0.34 | 0.101 | 0.10 | 0.634 |
| Avoid B, P(C) | -0.11 | 0.604 | 0.50 | **0.012** | 0.41 | 0.040 | 0.14 | 0.502 |
| Low Conflict, RT, ms | 0.13 | 0.547 | -0.27 | 0.195 | -0.30 | 0.148 | 0.18 | 0.394 |
| High Conflict, RT, ms | -0.22 | 0.290 | -0.42 | **0.034** | -0.47 | **0.019** | 0.25 | 0.226 |

##### 4.2.2. EEG and correlations with iron status

Supplementary Figure S7 presents the group averages for the response-locked (Supplementary Figure S7a) and feedback-locked (Supplementary Figure S7b) EEG features. Supplementary Table S17 presents the correlations involving all of the features and the iron status biomarkers. Each of the features was significantly related to iron status, such the higher iron status was related to larger (positive or negative) amplitudes. There were no significant correlations with any of the biomarkers associated with oxygen transport. Finally, age was significantly related with both of the FRNs, such that higher age was associated with lower amplitudes.

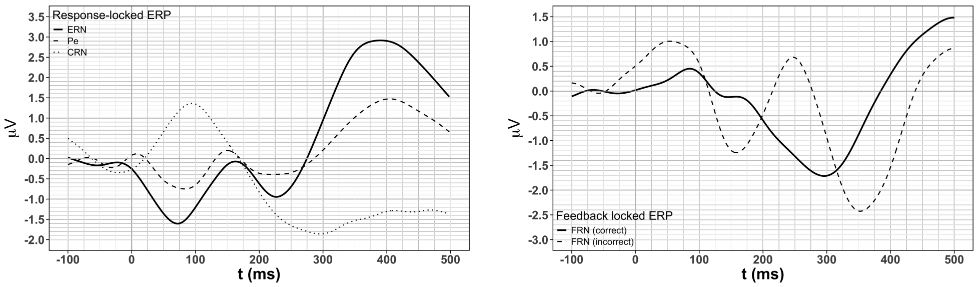

(a) (b)

**Figure S7.** Group average EEG data from the PST: (a) response-locked features, (b) feedback locked features.

**Table S17.** Correlations involving the EEG features from the PST with the iron status biomarkers. Cells highlighted in gray indicate correlations that were significant after correcting for the false discovery rate. Note: FRN (c) = FRN correct trials, FRN (i) = FRN incorrect trials

| | ERN | | Pe | | CRN | | FRN (c) | | FRN (i) | |
|---|---|---|---|---|---|---|---|---|---|---|
| | *r* | *p* | *r* | *p* | *r* | *p* | *r* | *p* | *r* | *p* |
| Age | 0.29 | 0.171 | -0.27 | 0.209 | 0.36 | 0.082 | 0.45 | **0.029** | 0.44 | **0.033** |
| Hb | -0.22 | 0.309 | 0.15 | 0.472 | -0.13 | 0.540 | -0.16 | 0.470 | -0.11 | 0.598 |
| sFt | -0.81 | **< 0.001** | 0.69 | **< 0.001** | -0.84 | **< 0.001** | -0.75 | **< 0.001** | -0.76 | **< 0.001** |
| sFt percentile | -0.88 | **< 0.001** | 0.73 | **< 0.001** | -0.94 | **< 0.001** | -0.91 | **< 0.001** | -0.92 | **< 0.001** |
| RBC | -0.13 | 0.537 | 0.23 | 0.284 | -0.02 | 0.922 | 0.02 | 0.940 | 0.01 | 0.960 |
| Hct | -0.11 | 0.618 | 0.06 | 0.780 | -0.05 | 0.829 | -0.04 | 0.863 | -0.01 | 0.950 |
| RDW | 0.35 | 0.092 | -0.33 | 0.113 | 0.25 | 0.236 | 0.25 | 0.236 | 0.27 | 0.198 |
| MCV | 0.05 | 0.823 | -0.28 | 0.194 | -0.05 | 0.815 | -0.09 | 0.661 | -0.05 | 0.830 |
| MCH | -0.13 | 0.560 | -0.10 | 0.633 | -0.17 | 0.427 | -0.25 | 0.242 | -0.18 | 0.404 |
| MCHC | -0.35 | 0.092 | 0.30 | 0.153 | -0.26 | 0.216 | -0.35 | 0.097 | -0.29 | 0.164 |

### *4.3. RBCL*

#### 4.3.1. Behavioral data

Descriptive statistics for each of the behavioral variables from the RBCL are presented in Supplementary Table S18. Each of the variables were analyzed using a one-way repeated measures mixed models ANOVA, with block (first 200 vs. second 200 trials) manipulated within participants and with participants as the random factor, and the results are presented in Supplementary Table S19. The analysis of the accuracy variable revealed a significant main effect of block, showing that participants performed more accurately in block 2 (M = .49, SE = .02) than in block 1 (M = .44, SE = .02), indicating a modest learning effect. Note that chance level of accuracy in this task is 0.25. This was the only variable for which there was a significant main effect for block.

**Table S18.** Descriptive statistics for the behavioral data from the RBCL.

| | Block 1 | | | | Block 2 | | | |
|---|---|---|---|---|---|---|---|---|
| Variable | M | SE | min | max | M | SE | min | max |
| Overall accuracy, P(C) | 0.44 | 0.02 | 0.30 | 0.71 | 0.49 | 0.02 | 0.29 | 0.71 |
| Marginal hit rate, orientation | 0.54 | 0.02 | 0.34 | 0.79 | 0.56 | 0.03 | 0.04 | 0.90 |
| Marginal false alarm rate, orientation | 0.36 | 0.03 | 0.09 | 0.67 | 0.27 | 0.03 | 0.10 | 0.61 |
| Marginal $d'$, orientation | 0.53 | 0.10 | -0.23 | 1.50 | 0.81 | 0.10 | -0.69 | 1.62 |
| Marginal $c$, orientation | 0.14 | 0.07 | -0.55 | 0.83 | 0.25 | 0.07 | -0.47 | 1.43 |
| Marginal hit rate, spatial frequency | 0.75 | 0.03 | 0.49 | 0.98 | 0.82 | 0.02 | 0.63 | 0.96 |
| Marginal false alarm rate, spatial frequency | 0.34 | 0.02 | 0.11 | 0.53 | 0.30 | 0.02 | 0.09 | 0.58 |
| Marginal $d'$, spatial frequency | 1.23 | 0.13 | 0.01 | 2.89 | 1.48 | 0.13 | 0.35 | 2.46 |
| Marginal $c$, spatial frequency | -0.17 | 0.05 | -0.75 | 0.30 | -0.17 | 0.05 | -0.73 | 0.59 |
| RT, correct responses, ms | 1059 | 27 | 837 | 1346 | 1024 | 27 | 798 | 1337 |

**Table S19.** Results of the repeated measures ANOVAs on the behavioral data from the RBCL.

| Variable | *F* | *p* |
|---|---|---|
| Overall accuracy, P(C) | 4.45 | 0.045 |
| Marginal $d'$, orientation | 4.24 | 0.050 |
| Marginal $c$, orientation | 1.32 | 0.261 |
| Marginal $d'$, spatial frequency | 2.95 | 0.098 |
| Marginal $c$, spatial frequency | 0.01 | 0.918 |
| RT, correct responses, ms | 2.29 | 0.143 |

Pearson correlations were run on accuracy and all of the other behavioral variables with the iron biomarkers and age, and the results are presented in Supplementary Table S20. No significant correlations involving accuracy were observed in block 1. However, there were significant positive correlations involving accuracy and sFt and sFt percentile in block 2, consistent with previous work with women of reproductive age [29,30].

**Table S20.** Correlations involving the behavioral data from the RBCL with the iron status biomarkers (1). Cells highlighted in gray indicate correlations that were statistically significant after correcting for the false discovery rate. Note: pctl = percentile.

| | | Hb | | sFt | | sFt pctl | | Age | |
|---|---|---|---|---|---|---|---|---|---|
| Variable | Block | *r* | *p* | *r* | *p* | *r* | *p* | *r* | *p* |
| Overall accuracy, P(C) | 1 | 0.06 | 0.758 | -0.14 | 0.486 | -0.15 | 0.826 | -0.23 | 0.267 |
| | 2 | -0.21 | 0.306 | 0.51 | **0.008** | 0.52 | **0.006** | -0.10 | 0.633 |
| Hit rate (%), orientation | 1 | 0.18 | 0.378 | -0.18 | 0.380 | -0.16 | 0.444 | 0.10 | 0.626 |
| | 2 | 0.07 | 0.735 | 0.50 | **0.010** | 0.42 | 0.035 | 0.04 | 0.843 |
| False alarm rate (%), orientation | 1 | -0.01 | 0.978 | 0.05 | 0.798 | 0.00 | 0.993 | 0.25 | 0.227 |
| | 2 | 0.22 | 0.291 | 0.11 | 0.593 | 0.03 | 0.100 | 0.09 | 0.669 |
| Marginal $d'$, orientation | 1 | 0.14 | 0.499 | -0.20 | 0.323 | -0.13 | 0.539 | -0.19 | 0.363 |
| | 2 | -0.13 | 0.534 | 0.37 | 0.062 | 0.37 | 0.060 | -0.06 | 0.764 |
| Marginal $c$, orientation | 1 | -0.08 | 0.687 | 0.04 | 0.859 | 0.07 | 0.742 | -0.23 | 0.266 |
| | 2 | -0.11 | 0.577 | -0.42 | 0.034 | -0.32 | 0.116 | -0.06 | 0.780 |
| Hit rate (%), spatial frequency | 1 | -0.12 | 0.565 | -0.03 | 0.887 | 0.07 | 0.752 | -0.25 | 0.222 |
| | 2 | 0.04 | 0.843 | 0.56 | **0.003** | 0.59 | **0.002** | -0.18 | 0.391 |
| False alarm rate (%), spatial frequency | 1 | 0.01 | 0.974 | 0.17 | 0.407 | 0.08 | 0.709 | 0.16 | 0.441 |
| | 2 | 0.28 | 0.159 | 0.13 | 0.526 | 0.01 | 0.967 | 0.25 | 0.223 |
| Marginal $d'$, spatial frequency | 1 | -0.13 | 0.521 | -0.17 | 0.395 | -0.07 | 0.794 | -0.24 | 0.228 |
| | 2 | -0.31 | 0.123 | 0.38 | 0.054 | 0.46 | **0.018** | -0.27 | 0.175 |
| Marginal $c$, spatial frequency | 1 | 0.21 | 0.311 | 0.01 | 0.969 | -0.04 | 0.851 | 0.18 | 0.379 |
| | 2 | -0.08 | 0.702 | -0.58 | **0.002** | -0.51 | **0.007** | -0.04 | 0.846 |
| RT, correct responses, ms | 1 | -0.03 | 0.870 | 0.20 | 0.336 | 0.08 | 0.700 | 0.35 | 0.077 |
| | 2 | 0.06 | 0.782 | -0.15 | 0.462 | -0.25 | 0.226 | 0.18 | 0.385 |

Pearson correlations of overall accuracy and the marginal signal detection measures, including marginal hit rate, marginal false alarm rate, marginal $d'$, and marginal $c$, with the iron status biomarkers and age were calculated for both blocks. Overall accuracy was positively correlated with sFt and sFt percentile in block 2, indicating that higher values of these iron biomarkers were associated with higher asymptotic levels of performance, consistent with work with women of reproductive age [29,30]. The marginal hit rate for orientation was also positively correlated with both sFt and sFt percentile in block 2, also consistent with results with women of reproductive age [29,30].

No significant correlations were found between the iron markers and age with marginal false alarm rate or marginal $d'$ for orientation in either block. There was, however, a negative correlation between marginal $c$ for orientation and sFt in block 2, showing that higher levels of sFt were associated with more liberal responding in the final portion of the task. While this last result is consistent with associations involving response criterion noted earlier, and with work with women of reproductive age [31], this suggests that iron levels were affecting only response bias and not sensitivity for orientation, an outcome which is inconsistent with previous work with this task [30].

The marginal hit rate for spatial frequency was positively correlated with both sFt and sFt percentile in block 2, indicating that higher hit rates for spatial frequency were associated with higher values of sFt and sFt percentile, which aligns with previous literature suggesting that higher levels of sFt are related to improvements in performance on this task [30]. No significant correlations were found between the iron markers with false alarms rate for spatial frequency in either block, which was unexpected due to the previous findings that lower sFt values predicted greater reductions in false alarm rates [32]. Marginal $d'$ for spatial frequency was positively correlated with sFt percentile in block 2, indicating that higher levels of sFt were associated with higher asymptotic discriminability for spatial frequency, a result that is consistent with prior results with women of reproductive age [29,30]. Marginal $c$ for spatial frequency was negatively correlated with both sFt and sFt percentile in block 2. This result is consistent with the results for orientation. However, sFt levels were associated with differences in both sensitivity and response bias, consistent with previous work with women of reproductive age [30].

Reaction times for correct responses were analyzed using a one-way repeated measures mixed models ANOVA, with block (first 200 vs. second 200 trials) manipulated within participants and participants as the random factor. Results did not indicate any significant main effects of RT for block. This is inconsistent with previous results with women of reproductive age, in which increasing practice with the task led to reductions in RT [30]. Since there was a main effect of accuracy but not RT, this may suggest that participants are prioritizing accuracy over speed, taking more time to respond to ensure correct answers, even if it results in slower response times.

Pearson correlations involving RT were conducted. No significant correlations were found for block 1. However, RT was positively correlated with RDW in block 2, indicating that longer RTs were associated with higher values of RDW. Since higher values of RDW are associated with poorer oxygen transport capability, this result is consistent with expectations.

##### 4.3.2. Correlations between EEG and iron status

The group average waveforms for the stimulus- and feedback locked EEG data are presented in Supplementary Figure S8. The complete set of correlations of these features with the iron status biomarkers are presented in Supplementary Table **??**. Note that these data are all collapsed across blocks in order to obtain the most stable estimates for correct and incorrect trials. Age was negatively correlated with the amplitudes of the P300 (both correct and incorrect) and the late positive slow wave (both correct and incorrect), indicating lower amplitudes with increasing age. The amplitudes of both of these features, for both correct and incorrect trials, were positively related to both sFt and sFt percentile, as was fractional area latency. This is consistent with previous work with women of reproductive age who were iron deficienct but not anemic [32,33] in a sample of women who were not iron deficient. The amplitude of the late positive slow wave, for both correct and incorrect trials, was negatively related to RDW, and the fractional area latency for correct trials was positively related to MCH. Both of these findings that improved iron status with respect to oxygen transport capability was related to better brain function, with this being true in a sample of women who were not anemic.

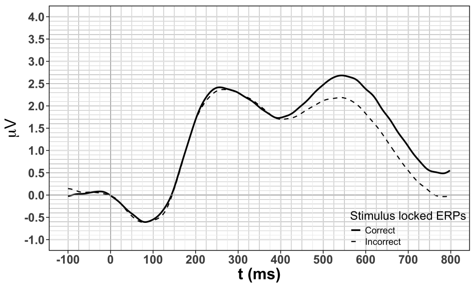

(a)

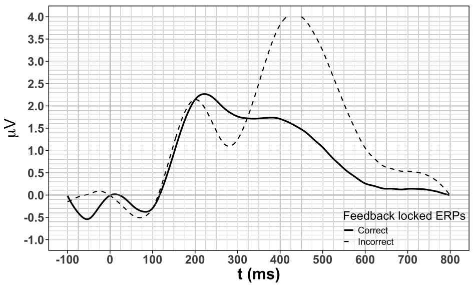

(b)

**Figure S8.** Group average EEG data from the RBCL: (a) stimulus-locked features, (b) feedback locked features.

#### *4.4. VSWM*

##### 4.4.1. Behavioral data

Descriptive statistics for all of the behavioral variables are presented in Supplementary Table S22. Accuracy and RT data were analyzed using 2 (number of targets: 3, 5) × 2 (number of distractors: 0, 2) × 2 (target: absent, present) repeated measures mixed models ANOVAs, with all three factors manipulated within participants and participants as the random factor. The signal detection measures ($d'$, $c$) and the working memory capacity measures ($K$, $K'$) were analyzed using 2 (number of targets: 3, 5) × 2 (number of distractors: 0, 2) repeated measures mixed models ANOVAs, with both factors manipulated within participants and participants as the random factor. The results of these analyses are presented in Supplementary Table S23.

**Table S22.** Descriptive statistics for the behavioral variables from the VSWM. Note: pres/abs = present/absent.

| | Targets | Distractors | Target pres/abs | M | SE | min | max |
|---|---|---|---|---|---|---|---|
| Accuracy P(C) | 3 | 0 | A | 0.970 | 0.03 | 0.83 | 1.00 |
| | | | P | 0.970 | 0.03 | 0.83 | 1.00 |
| | | 2 | A | 0.960 | 0.03 | 0.83 | 1.00 |
| | | | P | 0.960 | 0.03 | 0.67 | 1.00 |
| | 5 | 0 | A | 0.910 | 0.03 | 0.67 | 1.00 |
| | | | P | 0.800 | 0.03 | 0.00 | 1.00 |
| | | 2 | A | 0.880 | 0.03 | 0.58 | 1.00 |
| | | | P | 0.800 | 0.03 | 0.00 | 1.00 |
| RT, ms | 3 | 0 | A | 933 | 40 | 562 | 1238 |
| | | | P | 918 | 40 | 540 | 1324 |
| | | 2 | A | 941 | 40 | 581 | 1420 |
| | | | P | 915 | 40 | 606 | 1408 |
| | 5 | 0 | A | 989 | 40 | 614 | 1358 |
| | | | P | 984 | 40 | 563 | 1351 |
| | | 2 | A | 1051 | 40 | 591 | 1591 |
| | | | P | 992 | 40 | 704 | 1300 |
| Hit rate | 3 | 0 | | 0.96 | 0.01 | 0.83 | 0.99 |
| | | 2 | | 0.95 | 0.01 | 0.67 | 0.99 |
| | 5 | 0 | | 0.81 | 0.04 | 0.01 | 0.99 |
| | | 2 | | 0.80 | 0.05 | 0.01 | 0.99 |
| False alarm rate | 3 | 0 | | 0.04 | 0.01 | 0.01 | 0.17 |
| | | 2 | | 0.04 | 0.01 | 0.01 | 0.17 |
| | 5 | 0 | | 0.10 | 0.02 | 0.01 | 0.33 |
| | | 2 | | 0.12 | 0.02 | 0.01 | 0.42 |
| $d'$ | 3 | 0 | | 4.02 | 0.15 | 2.35 | 4.65 |
| | | 2 | | 3.94 | 0.19 | 1.40 | 4.65 |
| | 5 | 0 | | 2.65 | 0.26 | 0.00 | 4.65 |
| | | 2 | | 2.57 | 0.26 | 0.00 | 4.65 |
| $c$ | 3 | 0 | | 0.00 | 0.05 | -0.47 | 0.47 |
| | | 2 | | 0.00 | 0.05 | -0.47 | 0.47 |
| | 5 | 0 | | 0.27 | 0.11 | -0.47 | 2.33 |
| | | 2 | | 0.13 | 0.14 | -0.95 | 2.33 |
| $K$ | 3 | 0 | | 2.88 | 0.03 | 2.45 | 2.97 |
| | | 2 | | 2.84 | 0.05 | 1.80 | 2.97 |
| | 5 | 0 | | 3.88 | 0.23 | 0.00 | 4.95 |
| | | 2 | | 3.88 | 0.27 | 0.00 | 4.95 |
| $K'$ | 3 | 0 | | 0.40 | 0.03 | 0.38 | 0.43 |
| | | 2 | | 0.39 | 0.05 | 0.34 | 0.43 |
| | 5 | 0 | | 0.45 | 0.02 | 0.00 | 0.54 |
| | | 2 | | 0.43 | 0.03 | 0.00 | 0.54 |

**Table S23.** Repeated measures ANOVAs on the behavioral variables from the VSWM. Cells highlighted in gray indicate significant effects.

| Variable | Factor | *F* | *p* |
|---|---|---|---|
| Accuracy P(C) | Targets (T) | 32.56 | **< 0.001** |
| | Distractors (D) | 0.32 | 0.578 |
| | Present/absent (P) | 6.48 | **0.018** |
| | T x D | 0.03 | 0.875 |
| | T x P | 5.11 | **0.033** |
| | D x P | 0.06 | 0.809 |
| | T x D x P | 0.11 | 0.744 |
| RT, correct responses, ms | T | 27.19 | **< 0.001** |
| | D | 1.58 | 0.221 |
| | P | 3.20 | 0.086 |
| | T x D | 1.19 | 0.286 |
| | T x P | 0.17 | 0.682 |
| | D x P | 1.20 | 0.285 |
| | T x D x P | 0.53 | 0.474 |
| *K* | T | 25.22 | **< 0.001** |
| | D | 0.03 | 0.860 |
| | T x D | 0.01 | 0.920 |
| *K'* | T | 5.86 | **0.023** |
| | D | 0.46 | 0.505 |
| | T x D | 0.29 | 0.598 |
| *d'* | T | 46.73 | **< 0.001** |
| | D | 0.44 | 0.514 |
| | T x D | 0.00 | 0.956 |
| *c* | T | 3.69 | 0.066 |
| | D | 0.71 | 0.406 |
| | T x D | 0.51 | 0.480 |

Results for accuracy showed a main effect of number of targets, indicating that participants performed more accurately when 3 targets were present (M = .97, SE = .01) than when 5 targets were present (M = .85, SE = .01). This shows that accuracy was higher when there were fewer items to be remembered, which is to be expected [34]. Results also indicated that accuracy was higher when the target was absent (M = .93, SE = .01) relative to when it was present (M = .88, SE = .01), suggesting that participants were more accurate at rejecting a probe at a location that was not occupied by a target than they were at confirming a probe at a location that was occupied by a target, which also aligns with previous research [34]. There was a significant interaction between the number of targets presented and whether a target was present or absent, wherein participants were more accurate when the target was absent but that this was true only when five targets were presented (Supplementary Figure S9). This could indicate that the five target condition was demanding to the point of there being no advantage for target absent over target present trials.

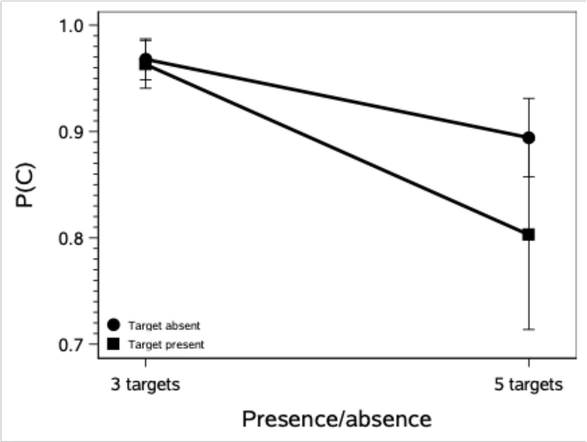

**Figure S9.** Interaction of number of targets and target present/absent in the accuracy data, VSWM.

Results of the analyses of the RT data revealed a main effect of number of targets, indicating that participants responded faster when three targets were presented (M = 927, SE = 35) vs. when five targets were presented (M = 1004, SE = 35). This is consistent with the regularity that lower set sizes are associated with shorter RTs in healthy adults [35,36]. None of the other effects were significant.

Two measures of working memory capacity were calculated. The first [34], $K$, was calculated as

$$K = N \times (H - F)$$

where $N$ is the number of targets to be remembered, $H$ is the hit rate, and $F$ is the false alarm rate. The form of this measure guarantees that $K$ will always be higher for five targets than for three targets. To adjust for this and provide a more interpretable measure across levels of total memory load, we also calculated a second capacity measure, $K'$, which we defined as

$$K' = \frac{N \times (H - F)}{N + D}$$

where $D$ is the number of distractors.

The repeated measures ANOVA on $K$ revealed a main effect of number of targets, showing that participants had a higher working memory capacity when five targets were presented relative to when three targets were presented (M = 2.86, SE = 0.04). As noted above, this is a natural consequence of the way that $K$ is defined. The repeated measures ANOVA was on $K'$ also found a higher working memory capacity when five targets were presented (M = .44 , SE = .01) relative to when three targets were presented (M = .40, SE = .01). The magnitude of the difference, however, suggests that while the difference was significant, it most likely is not meaningful.

The repeated measures ANOVA on $d'$ revealed a main effect of number of targets, showing that participants had a higher $d'$ when three targets were presented (M = 3.98, SE = .15) vs. when five targets were presented (M = 2.61, SE = .23). This shows that the participants found it easier to differentiate between the presence and absence of a target when the three targets were presented, which is consistent with classic findings in the literature [37]. There were no significant effects on $c$.

##### 4.4.2. Correlations between behavior and iron status

The complete set of correlations involving the behavioral variables and the iron status biomarkers and age are presented in Supplementary Tables **??**-S26. Age was negatively correlated with accuracy, but only when five targets and zero distractors were present. Age was positively correlated with RT in all conditions, indicating age-related slowing of performance [38]. Finally, age was positively correlated with the false alarm rate, but only when five targets and zero distractors were present.

**Table S26.** Correlations involving the behavioral variables from the VSWM and the iron status biomarkers (3). Cells highlighted in gray indicate correlations that were signficant after correcting for the false discovery rate.

| Variable | Targets | Distractors | Target pres/abs | MCHC | | RDW | |
|---|---|---|---|---|---|---|---|
| | | | | *r* | *p* | *r* | *p* |
| Accuracy P(C) | 3 | 0 | A | -0.24 | 0.229 | 0.21 | 0.307 |
| | | | P | 0.21 | 0.292 | -0.09 | 0.651 |
| | | 2 | A | -0.13 | 0.534 | 0.26 | 0.192 |
| | | | P | -0.17 | 0.409 | 0.14 | 0.484 |
| | 5 | 0 | A | -0.04 | 0.847 | -0.11 | 0.577 |
| | | | P | -0.02 | 0.919 | 0.00 | 0.990 |
| | | 2 | A | -0.07 | 0.740 | 0.13 | 0.533 |
| | | | P | 0.05 | 0.794 | -0.41 | 0.040 |
| RT, correct responses | 3 | 0 | A | 0.00 | 0.981 | 0.31 | 0.126 |
| | | | P | -0.03 | 0.877 | 0.36 | 0.068 |
| | | 2 | A | -0.03 | 0.894 | 0.37 | 0.064 |
| | | | P | -0.03 | 0.883 | 0.31 | 0.123 |
| | 5 | 0 | A | -0.11 | 0.590 | 0.32 | 0.106 |
| | | | P | 0.01 | 0.953 | 0.41 | 0.045 |
| | | 2 | A | 0.04 | 0.850 | 0.27 | 0.185 |
| | | | P | -0.06 | 0.759 | 0.41 | 0.039 |
| Hit rate | 3 | 0 | | 0.21 | 0.303 | -0.09 | 0.679 |
| | | 2 | | -0.17 | 0.396 | 0.15 | 0.471 |
| | 5 | 0 | | -0.03 | 0.870 | 0.00 | 0.982 |
| | | 2 | | 0.05 | 0.793 | -0.40 | 0.042 |
| False alarm rate | 3 | 0 | | 0.25 | 0.226 | -0.21 | 0.297 |
| | | 2 | | 0.14 | 0.494 | -0.27 | 0.190 |
| | 5 | 0 | | 0.04 | 0.851 | 0.17 | 0.573 |
| | | 2 | | 0.06 | 0.779 | -0.12 | 0.546 |
| *K* | 3 | 0 | | 0.19 | 0.358 | -0.06 | 0.760 |
| | | 2 | | -0.18 | 0.366 | 0.16 | 0.440 |
| | 5 | 0 | | -0.08 | 0.709 | 0.00 | 0.984 |
| | | 2 | | 0.04 | 0.851 | -0.39 | 0.051 |
| $K'$ | 3 | 0 | | -0.03 | 0.902 | 0.04 | 0.856 |
| | | 2 | | -0.19 | 0.353 | 0.06 | 0.763 |
| | 5 | 0 | | -0.10 | 0.618 | 0.07 | 0.721 |
| | | 2 | | 0.06 | 0.769 | -0.35 | 0.078 |
| $d'$ | 3 | 0 | | 0.01 | 0.960 | 0.03 | 0.897 |
| | | 2 | | -0.11 | 0.608 | 0.19 | 0.347 |
| | 5 | 0 | | 0.00 | 0.998 | -0.12 | 0.568 |
| | | 2 | | -0.05 | 0.800 | -0.35 | 0.084 |
| *c* | 3 | 0 | | -0.41 | 0.036 | 0.28 | 0.172 |
| | | 2 | | 0.07 | 0.722 | 0.14 | 0.493 |
| | 5 | 0 | | -0.05 | 0.820 | 0.00 | 0.984 |
| | | 2 | | -0.12 | 0.552 | 0.45 | **0.022** |

Accuracy, hit rates, working memory capacity ($K$ and $K'$), and $d'$ were all positively related to sFt and were more frequently positively related to sFt percentile. This indicates that numerous measures of performance were related to these measures of iron status in the same way as has been observed in women of reproductive age. In addition, higher values of both of these were related to more liberal responding, something that we have observed in previous studies with women of reproductive age. With respect to Hb and the other iron status variables related to oxygen transport, the only correlation to reach significance after correcting for the false discovery rate was a positive relationship between $c$ and RDW, indicating that a lower iron status (higher values of RDW) were associated with a more conservative response, something that we have observed in previous studies involving women of reproductive age.

##### 4.4.3. EEG data

Group average waveforms at each level of number of targets and number of distractors at each of the three electrodes are presented in the panels of Supplementary Figure S10. These data were analyzed separately at each electrode using a set of 2 (number of targets: 3, 5) $\times$ 2 (number of distractors) $\times$ 2 (match: match, non-match) mixed models repeated measures ANOVA, with the three factors as fixed and participant as a random factor. In this section of the results "match" refers to a target present trial and "non-match" refers to a target absent trial. The results of these analysis are presented in Supplementary Table S27. None of the main effects or interactions were significant.

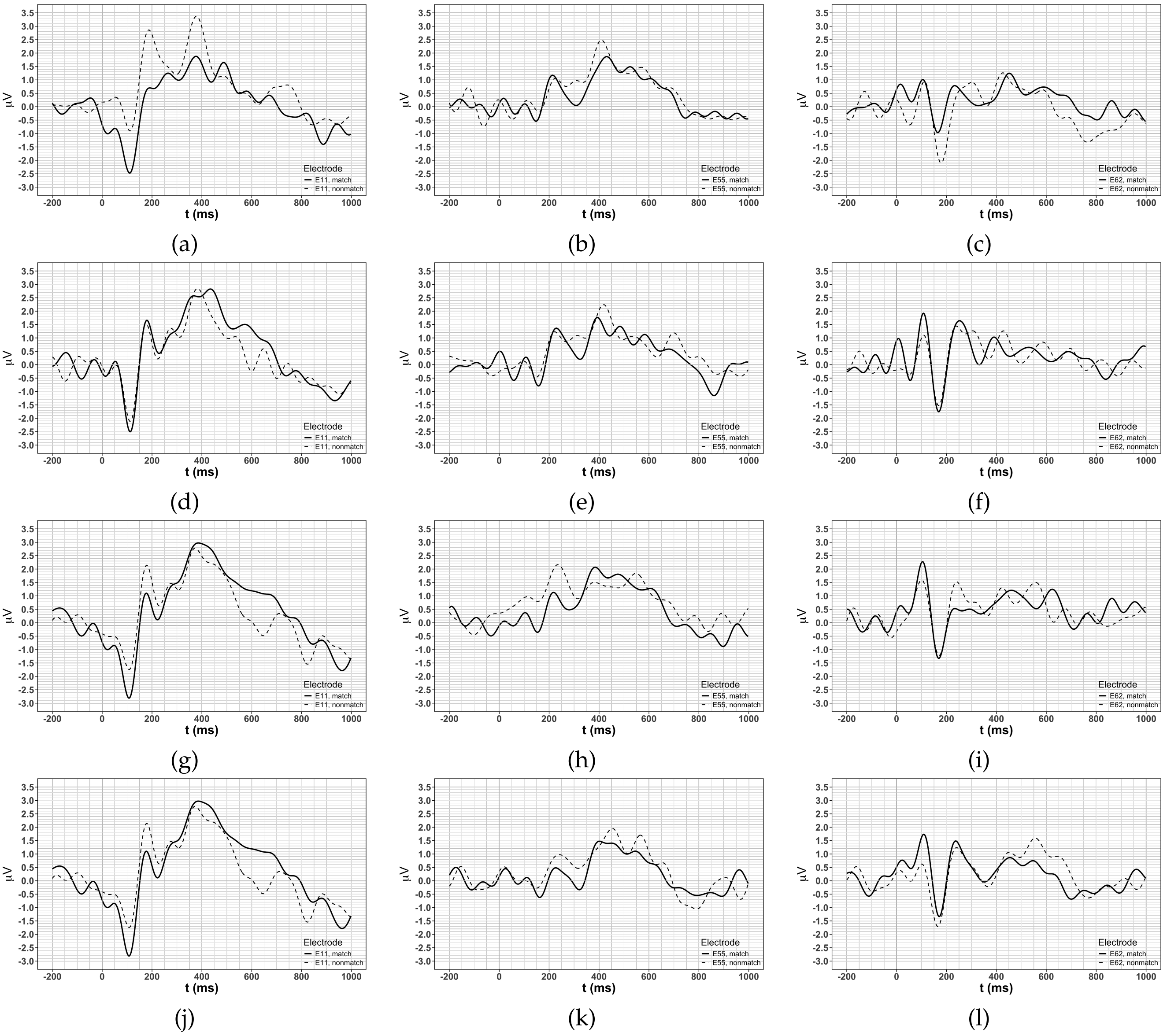

**Figure S10.** Group average waveforms at each level of number of targets and number of distractors at each of the three electrode: (a)-(c), 3 targets, 0 distractors; (d)-(f), 3 targets, 2 distractors; (g)-(i), 5 targets, 0 distractors; (j)-(l) 5 targets, 2 distractors. Solid lines are from target present trials and dashed lines are from target absent trials.

**Table S27.** Repeated measures analyses of the EEG data from the VSWM.

| Electrode | Factor | $F$ | $p$ |
|---|---|---|---|
| E11 | Targets (T) | 2.40 | 0.124 |
| | Distractors (D) | 0.40 | 0.527 |
| | Match (M) | 3.26 | 0.073 |
| | T × D | 0.01 | 0.907 |
| | T × M | 0.42 | 0.520 |
| | D × $M$ | 0.01 | 0.927 |
| | T × D × M | 1.68 | 0.197 |
| E55 | T | 0.45 | 0.501 |
| | D | 3.30 | 0.071 |
| | M | 2.12 | 0.148 |
| | T × D | 2.16 | 0.144 |
| | T × M | 0.00 | 0.963 |
| | D × M | 0.11 | 0.743 |
| | T × D × M | 0.06 | 0.810 |
| E62 | T | 0.14 | 0.708 |
| | D | 0.01 | 0.920 |
| | M | 0.45 | 0.505 |
| | T × D | 0.70 | 0.405 |
| | T × M | 0.01 | 0.940 |
| | D × M | 0.01 | 0.941 |
| | T × D × M | 0.39 | 0.531 |

##### 4.4.4. VSWM EEG correlations

The complete set of correlations involving the EEG variables from the VSWM and the iron status biomarkers and age are presented in Supplementary Tables S28-S30. The only correlation involving age was for amplitudes at electrode E11 on trials on which three targets and 0 distractors. That relationship was positive, which runs counter to expectations. There were a small number of positive correlations between amplitudes and sFt, with these indicating that higher sFt values were associated with larger amplitudes, a result that is consistent with prior work with women of reproductive age [31,33]. There were no significant relationships between amplitudes at any of the electrodes and sFt percentile. There were no significant correlations involving Hb. However, there were a small number of significant correlations involving amplitude and either MCH and MCHC, indicating that there was some evidence for a positive relationship between these iron status biomarkers—related to oxygen transport—and brain function.

**Table S28.** Correlations involving the EEG variables from the VSWM from the iron status biomarkers (1). Cells highlighted in gray indicate correlations that were significant after correcting for the false discovery rate.

| Targets | Distractors | Match | Variable | Electrode | *r* | *p* | Targets | Distractors | Match | *r* | *p* |
|---|---|---|---|---|---|---|---|---|---|---|---|
| 3 | 0 | 0 | Age | E11 | 0.49 | **0.021** | 5 | 0 | 0 | 0.30 | 0.180 |
| | | | | E55 | 0.14 | 0.531 | | | | 0.16 | 0.482 |
| | | | | E62 | 0.06 | 0.777 | | | | 0.29 | 0.198 |
| | | | Hb | E11 | 0.12 | 0.607 | | | | 0.13 | 0.574 |
| | | | | E55 | -0.22 | 0.329 | | | | -0.10 | 0.664 |
| | | | | E62 | -0.22 | 0.324 | | | | -0.18 | 0.415 |
| | | | sFt | E11 | 0.22 | 0.315 | | | | 0.33 | 0.128 |
| | | | | E55 | 0.36 | 0.104 | | | | -0.03 | 0.882 |
| | | | | E62 | 0.10 | 0.644 | | | | -0.15 | 0.509 |
| | | | sFt_pctl | E11 | 0.04 | 0.871 | | | | 0.13 | 0.565 |
| | | | | E55 | 0.32 | 0.149 | | | | -0.10 | 0.646 |
| | | | | E62 | 0.13 | 0.564 | | | | -0.24 | 0.286 |
| | | 1 | Age | E11 | 0.22 | 0.322 | | | 1 | 0.24 | 0.287 |
| | | | | E55 | -0.27 | 0.227 | | | | 0.14 | 0.533 |
| | | | | E62 | -0.04 | 0.843 | | | | 0.18 | 0.428 |
| | | | Hb | E11 | -0.03 | 0.891 | | | | 0.13 | 0.571 |
| | | | | E55 | 0.33 | 0.134 | | | | -0.24 | 0.274 |
| | | | | E62 | 0.09 | 0.688 | | | | -0.19 | 0.401 |
| | | | sFt | E11 | 0.58 | **0.004** | | | | 0.45 | 0.034 |
| | | | | E55 | 0.34 | 0.118 | | | | 0.12 | 0.596 |
| | | | | E62 | -0.04 | 0.855 | | | | -0.19 | 0.396 |
| | | | sFt_pctl | E11 | 0.43 | 0.048 | | | | 0.33 | 0.134 |
| | | | | E55 | 0.45 | 0.036 | | | | 0.07 | 0.768 |
| | | | | E62 | 0.04 | 0.855 | | | | -0.28 | 0.199 |
| | 2 | 0 | Age | E11 | 0.18 | 0.410 | | 2 | 0 | 0.04 | 0.861 |
| | | | | E55 | 0.01 | 0.972 | | | | 0.13 | 0.568 |
| | | | | E62 | -0.08 | 0.726 | | | | 0.15 | 0.496 |
| | | | Hb | E11 | 0.08 | 0.725 | | | | 0.00 | 0.996 |
| | | | | E55 | 0.21 | 0.341 | | | | -0.26 | 0.237 |
| | | | | E62 | -0.01 | 0.976 | | | | -0.10 | 0.662 |
| | | | sFt | E11 | 0.48 | **0.023** | | | | 0.26 | 0.250 |
| | | | | E55 | 0.00 | 0.983 | | | | 0.01 | 0.981 |
| | | | | E62 | -0.08 | 0.735 | | | | -0.04 | 0.873 |
| | | | sFt_pctl | E11 | 0.38 | 0.079 | | | | 0.26 | 0.236 |
| | | | | E55 | -0.05 | 0.820 | | | | -0.06 | 0.808 |
| | | | | E62 | -0.01 | 0.973 | | | | -0.12 | 0.596 |
| | | 1 | Age | E11 | 0.10 | 0.665 | | | 1 | 0.12 | 0.601 |
| | | | | E55 | -0.02 | 0.922 | | | | 0.56 | **0.007** |
| | | | | E62 | 0.08 | 0.725 | | | | 0.42 | 0.054 |
| | | | Hb | E11 | 0.21 | 0.339 | | | | 0.02 | 0.921 |
| | | | | E55 | -0.25 | 0.266 | | | | -0.05 | 0.841 |
| | | | | E62 | -0.18 | 0.423 | | | | 0.11 | 0.623 |
| | | | sFt | E11 | 0.48 | **0.025** | | | | 0.51 | **0.015** |
| | | | | E55 | 0.04 | 0.856 | | | | 0.34 | 0.122 |
| | | | | E62 | -0.10 | 0.670 | | | | 0.23 | 0.312 |
| | | | sFt_pctl | E11 | 0.42 | 0.054 | | | | 0.40 | 0.069 |
| | | | | E55 | 0.03 | 0.879 | | | | 0.06 | 0.793 |
| | | | | E62 | -0.11 | 0.613 | | | | -0.01 | 0.973 |

**Table S29.** Correlations involving the EEG variables from the VSWM from the iron status biomarkers (2). Cells highlighted in gray indicate correlations that were significant after correcting for the false discovery rate.

| Targets | Distractors | Match | Variable | Electrode | *r* | *p* | Targets | Distractors | Match | *r* | *p* |
|---|---|---|---|---|---|---|---|---|---|---|---|
| 3 | 0 | 0 | RBC | E11 | -0.01 | 0.978 | 5 | 0 | 0 | 0.05 | 0.815 |
| | | | | E55 | -0.07 | 0.750 | | | | 0.03 | 0.889 |
| | | | | E62 | -0.08 | 0.720 | | | | -0.20 | 0.369 |
| | | | HCT | E11 | -0.03 | 0.904 | | | | 0.04 | 0.852 |
| | | | | E55 | -0.25 | 0.258 | | | | -0.14 | 0.537 |
| | | | | E62 | -0.21 | 0.339 | | | | -0.20 | 0.371 |
| | | | RDW | E11 | -0.07 | 0.770 | | | | -0.02 | 0.918 |
| | | | | E55 | -0.10 | 0.673 | | | | 0.00 | 0.984 |
| | | | | E62 | -0.27 | 0.223 | | | | 0.02 | 0.929 |
| | | 1 | RBC | E11 | -0.08 | 0.733 | | | 1 | 0.09 | 0.700 |
| | | | | E55 | 0.07 | 0.773 | | | | -0.14 | 0.533 |
| | | | | E62 | -0.26 | 0.250 | | | | -0.16 | 0.485 |
| | | | HCT | E11 | -0.11 | 0.611 | | | | 0.04 | 0.854 |
| | | | | E55 | 0.21 | 0.358 | | | | -0.26 | 0.241 |
| | | | | E62 | -0.02 | 0.919 | | | | -0.11 | 0.640 |
| | | | RDW | E11 | -0.14 | 0.531 | | | | -0.14 | 0.526 |
| | | | | E55 | -0.32 | 0.145 | | | | 0.08 | 0.717 |
| | | | | E62 | -0.34 | 0.123 | | | | 0.23 | 0.295 |
| | 2 | 0 | RBC | E11 | -0.22 | 0.319 | | 2 | 0 | -0.33 | 0.132 |
| | | | | E55 | 0.20 | 0.365 | | | | -0.25 | 0.263 |
| | | | | E62 | -0.01 | 0.950 | | | | -0.04 | 0.877 |
| | | | HCT | E11 | -0.06 | 0.807 | | | | -0.19 | 0.390 |
| | | | | E55 | 0.16 | 0.485 | | | | -0.13 | 0.571 |
| | | | | E62 | -0.11 | 0.634 | | | | 0.08 | 0.720 |
| | | | RDW | E11 | -0.27 | 0.216 | | | | -0.23 | 0.295 |
| | | | | E55 | 0.04 | 0.876 | | | | 0.21 | 0.341 |
| | | | | E62 | -0.24 | 0.288 | | | | 0.23 | 0.294 |
| | | 1 | RBC | E11 | 0.05 | 0.834 | | | 1 | -0.16 | 0.482 |
| | | | | E55 | -0.27 | 0.230 | | | | 0.23 | 0.307 |
| | | | | E62 | -0.31 | 0.160 | | | | 0.10 | 0.661 |
| | | | HCT | E11 | 0.10 | 0.657 | | | | -0.11 | 0.617 |
| | | | | E55 | -0.31 | 0.162 | | | | 0.08 | 0.724 |
| | | | | E62 | -0.24 | 0.272 | | | | 0.20 | 0.376 |
| | | | RDW | E11 | -0.13 | 0.572 | | | | -0.06 | 0.775 |
| | | | | E55 | -0.03 | 0.884 | | | | 0.19 | 0.400 |
| | | | | E62 | 0.05 | 0.836 | | | | -0.08 | 0.721 |

**Table S30.** Correlations involving the EEG variables from the VSWM from the iron status biomarkers (2). Cells highlighted in gray indicate correlations that were significant after correcting for the false discovery rate.

| Targets | Distractors | Match | Variable | Electrode | *r* | *p* | Targets | Distractors | Match | *r* | *p* |
|---|---|---|---|---|---|---|---|---|---|---|---|
| 3 | 0 | 0 | MCV | E11 | -0.03 | 0.901 | 5 | 0 | 0 | -0.02 | 0.932 |
| | | | | E55 | -0.23 | 0.304 | | | | -0.24 | 0.274 |
| | | | | E62 | -0.16 | 0.476 | | | | 0.05 | 0.830 |
| | | | MCH | E11 | 0.16 | 0.465 | | | | 0.10 | 0.643 |
| | | | | E55 | -0.20 | 0.363 | | | | -0.19 | 0.398 |
| | | | | E62 | -0.19 | 0.407 | | | | 0.03 | 0.893 |
| | | | MCHC | E11 | 0.43 | 0.045 | | | | 0.27 | 0.223 |
| | | | | E55 | 0.02 | 0.924 | | | | 0.06 | 0.775 |
| | | | | E62 | -0.09 | 0.690 | | | | -0.03 | 0.901 |
| | | 1 | MCV | E11 | -0.03 | 0.901 | | | 1 | -0.07 | 0.747 |
| | | | | E55 | 0.15 | 0.499 | | | | -0.14 | 0.549 |
| | | | | E62 | 0.35 | 0.106 | | | | 0.11 | 0.635 |
| | | | MCH | E11 | 0.07 | 0.744 | | | | 0.06 | 0.774 |
| | | | | E55 | 0.34 | 0.127 | | | | -0.14 | 0.527 |
| | | | | E62 | 0.47 | **0.027** | | | | -0.04 | 0.870 |
| | | | MCHC | E11 | 0.23 | 0.306 | | | | 0.30 | 0.180 |
| | | | | E55 | 0.47 | **0.027** | | | | -0.04 | 0.846 |
| | | | | E62 | 0.34 | 0.123 | | | | -0.29 | 0.183 |
| | 2 | 0 | MCV | E11 | 0.28 | 0.214 | | 2 | 0 | 0.27 | 0.232 |
| | | | | E55 | -0.12 | 0.601 | | | | 0.21 | 0.342 |
| | | | | E62 | -0.11 | 0.619 | | | | 0.16 | 0.477 |
| | | | MCH | E11 | 0.42 | 0.049 | | | | 0.47 | **0.027** |
| | | | | E55 | -0.01 | 0.966 | | | | -0.01 | 0.958 |
| | | | | E62 | 0.01 | 0.953 | | | | -0.08 | 0.716 |
| | | | MCHC | E11 | 0.40 | 0.067 | | | | 0.52 | **0.013** |
| | | | | E55 | 0.23 | 0.313 | | | | 0.46 | 0.033 |
| | | | | E62 | 0.26 | 0.243 | | | | 0.51 | **0.016** |
| | | 1 | MCV | E11 | 0.07 | 0.773 | | | 1 | 0.10 | 0.667 |
| | | | | E55 | -0.01 | 0.968 | | | | -0.27 | 0.233 |
| | | | | E62 | 0.15 | 0.519 | | | | 0.08 | 0.736 |
| | | | MCH | E11 | 0.23 | 0.294 | | | | 0.25 | 0.258 |
| | | | | E55 | 0.02 | 0.937 | | | | -0.39 | 0.076 |
| | | | | E62 | 0.17 | 0.452 | | | | -0.02 | 0.947 |
| | | | MCHC | E11 | 0.40 | 0.066 | | | | 0.38 | 0.081 |
| | | | | E55 | 0.06 | 0.779 | | | | -0.33 | 0.134 |
| | | | | E62 | 0.34 | 0.123 | | | | -0.18 | 0.433 |

### *4.5. Blink rates*

The complete set of correlations between task-related and spontaneous blink rates (from the resting period) are presented in Supplementary Table S31. There were no significant correlations with age. Task-related blink rates in each of the tasks were all significantly negatively related to blink rates, with this consistent with the idea that greater cognitive engagement in a task is related to lower blink rates [39,40]. Task-related blink rates in both the FNAM and RBCL were negatively related to blink rates, and task-related blink rates in the FNAM, PST, and RBCL, were negatively related to at least one other biomarker associated with oxygen transport.

**Table S31.** Correlations involving task-related and spontaneous blink rates and the iron status biomarkers and age. Cells highlighted in gray indicate correlations that were significant after correcting for the false discovery rate.

| | FNAM | | PST | | RBCL | | VSWM | | Resting | |
|---|---|---|---|---|---|---|---|---|---|---|
| | $r$ | $p$ | $r$ | $p$ | $r$ | $p$ | $r$ | $p$ | $r$ | $p$ |
| Age | 0.21 | 0.323 | 0.15 | 0.465 | 0.15 | 0.47 | 0.09 | 0.668 | 0.00 | 0.986 |
| Hb | -0.49 | **0.016** | -0.25 | 0.217 | -0.50 | **0.011** | -0.36 | 0.081 | 0.00 | 0.986 |
| sFt | -0.57 | **0.004** | -0.47 | **0.017** | -0.43 | **0.032** | -0.53 | **0.006** | -0.34 | 0.108 |
| sFt percentile | -0.60 | **0.002** | -0.47 | **0.015** | -0.45 | **0.025** | -0.54 | **0.006** | -0.38 | 0.077 |
| RBC | -0.24 | 0.623 | -0.22 | 0.368 | -0.39 | 0.056 | -0.31 | 0.138 | -0.11 | 0.626 |
| Hct | -0.41 | 0.062 | -0.20 | **0.032** | -0.37 | 0.068 | -0.36 | 0.079 | 0.05 | 0.815 |
| RDW | 0.51 | **0.007** | 0.21 | 0.050 | 0.29 | 0.154 | 0.24 | 0.253 | 0.11 | 0.611 |
| MCV | -0.33 | 0.470 | -0.27 | 0.751 | 0.13 | 0.551 | 0.00 | 0.985 | 0.21 | 0.330 |
| MCH | -0.39 | 0.132 | -0.33 | 0.816 | -0.11 | 0.597 | -0.05 | 0.797 | 0.13 | 0.561 |
| MCHC | -0.19 | 0.083 | -0.18 | 0.276 | -0.47 | **0.017** | -0.13 | 0.528 | -0.14 | 0.529 |

#### *4.6. MRI estimates of brain iron deposits*

Initial inspection of the intensity values from each of the ROIs indicated that the data were not normally distributed and that variances were quite different across ROIs. Therefore, the intensity values were transformed using the Box-Cox transformation

$$y' = \frac{y^{\lambda} - 1}{\lambda}, y \neq 0$$

with $\lambda = 0.22$. Group averages for transformed intensity values in each ROI in each hemisphere are presented in Supplementary Figure S11. As a reminder, lower intensity values are indicative of higher iron deposits. These data were analyzed using a 2 (hemisphere: left, right) × 6 (ROI: caudate, putamen, globus pallidus, substantia nigra, VLPFC, ACC) mixed model repeated measures ANOVA, with hemisphere and ROI as fixed factors and participant as the random factor. The results of this analysis are presented in Supplementary Table S32. There was a significant main effect of hemisphere and a significant hemisphere × ROI interaction, with post-hoc Tukey tests indicating that intensity values were lowest in substantia nigra, globus pallidus, caudate, and putamen being lower (higher in iron) than VLPFC which in turn was lower than ACC, with the ordering of the lowest values being different in each hemisphere. These results are consistent with previous literature on brain iron deposits [27,28,41,42].

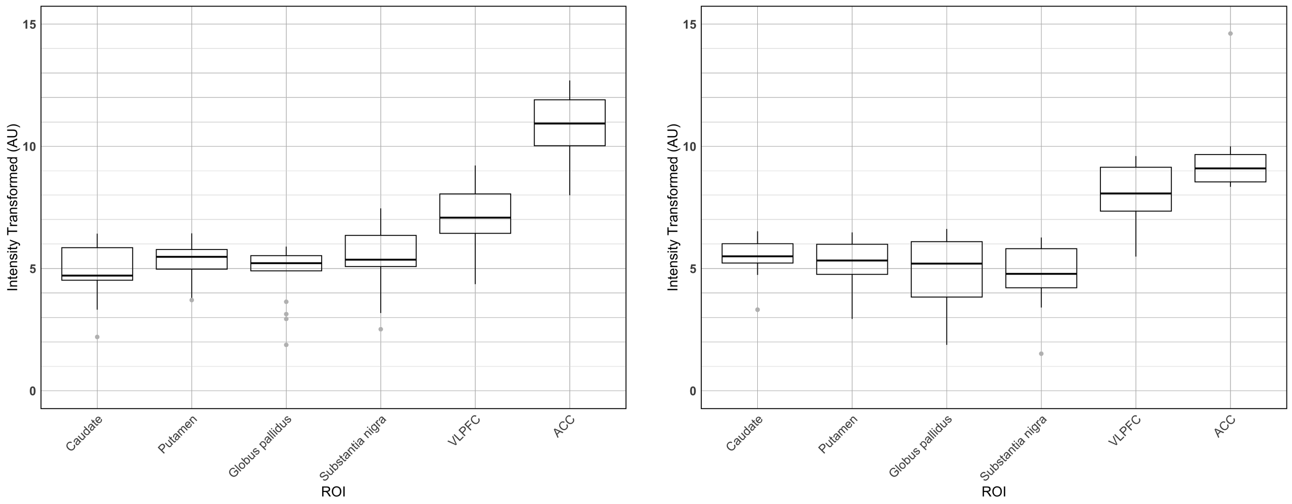

(a) (b)

**Figure S11.** Transformed intensity values for each of the ROIs in (a) the left and (b) the right hemisphere. Note: AU = arbitrary units.

**Table S32.** Repeated measures analysis of the transformed MRI intensity values.

| Factor | *df* | *F* | *p* |
|---|---|---|---|
| Hemisphere (H) | 1 | 0.35 | 0.556 |
| ROI (R) | 5 | 103.79 | < 0.001 |
| H × R | 5 | 3.79 | 0.003 |

The complete set of correlations involving the MRI estimates of brain iron deposits and the blood iron status biomarkers are presented in Supplementary Tables S33-S35. In brief, there were no significant correlations observed.

**Table S33.** Correlations involving transformed MRI intensity values and blood iron biomarkers (1).

| | | Age | | Hb | | sFt | | sFt pctl | |
|---|---|---|---|---|---|---|---|---|---|
| Hemisphere | ROI | *r* | *p* | *r* | *p* | *r* | *p* | *r* | *p* |
| Left | Caudate | -0.07 | 0.794 | -0.19 | 0.470 | 0.20 | 0.443 | 0.18 | 0.496 |
| | Putamen | -0.32 | 0.217 | 0.22 | 0.401 | -0.26 | 0.317 | -0.11 | 0.673 |
| | Globus pallidus | 0.21 | 0.426 | -0.35 | 0.173 | -0.32 | 0.208 | -0.36 | 0.161 |
| | Substantia nigra | -0.01 | 0.966 | 0.17 | 0.519 | -0.15 | 0.578 | -0.16 | 0.536 |
| | VLPFC | 0.30 | 0.247 | 0.20 | 0.444 | -0.25 | 0.335 | -0.28 | 0.269 |
| | ACC | 0.28 | 0.286 | 0.04 | 0.877 | -0.13 | 0.633 | -0.30 | 0.240 |
| Right | Caudate | 0.05 | 0.860 | 0.18 | 0.482 | -0.23 | 0.369 | -0.24 | 0.358 |
| | Putamen | -0.23 | 0.366 | -0.02 | 0.942 | 0.12 | 0.646 | 0.15 | 0.575 |
| | Globus pallidus | 0.28 | 0.269 | 0.19 | 0.466 | -0.20 | 0.439 | -0.27 | 0.287 |
| | Substantia nigra | 0.11 | 0.691 | -0.21 | 0.446 | -0.30 | 0.262 | -0.36 | 0.172 |
| | VLPFC | 0.11 | 0.663 | 0.35 | 0.164 | -0.29 | 0.259 | -0.27 | 0.293 |
| | ACC | -0.09 | 0.724 | -0.12 | 0.661 | -0.32 | 0.211 | -0.49 | 0.045 |

**Table S34.** Correlations involving transformed MRI intensity values and blood iron biomarkers (2).

| | | RBC | | HCT | | RDW | |
|---|---|---|---|---|---|---|---|
| Hemisphere | ROI | *r* | *p* | *r* | *p* | *r* | *p* |
| Left | Caudate | 0.02 | 0.943 | -0.34 | 0.183 | -0.15 | 0.571 |
| | Putamen | -0.08 | 0.757 | 0.29 | 0.252 | -0.21 | 0.411 |
| | Globus pallidus | -0.11 | 0.680 | -0.19 | 0.476 | 0.41 | 0.103 |
| | Substantia nigra | 0.30 | 0.236 | 0.09 | 0.746 | 0.37 | 0.142 |
| | VLPFC | 0.07 | 0.790 | 0.32 | 0.217 | -0.12 | 0.655 |
| | ACC | -0.02 | 0.955 | 0.21 | 0.422 | 0.05 | 0.865 |
| Right | Caudate | 0.41 | 0.105 | 0.07 | 0.789 | 0.12 | 0.659 |
| | Putamen | -0.31 | 0.230 | -0.10 | 0.705 | -0.15 | 0.572 |
| | Globus pallidus | 0.34 | 0.179 | 0.21 | 0.428 | 0.15 | 0.580 |
| | Substantia nigra | -0.01 | 0.970 | -0.25 | 0.342 | -0.33 | 0.209 |
| | VLPFC | 0.25 | 0.326 | 0.49 | 0.047 | -0.21 | 0.430 |
| | ACC | 0.40 | 0.117 | 0.36 | 0.155 | -0.13 | 0.613 |

**Table S35.** Correlations involving transformed MRI intensity values and blood iron biomarkers (3).

| | | MCV | | MCH | | MCHC | |
|---|---|---|---|---|---|---|---|
| Hemisphere | ROI | *r* | *p* | *r* | *p* | *r* | *p* |
| Left | Caudate | -0.41 | 0.100 | -0.27 | 0.299 | 0.21 | 0.417 |
| | Putamen | 0.44 | 0.080 | 0.36 | 0.153 | -0.04 | 0.880 |
| | Globus pallidus | -0.08 | 0.768 | -0.33 | 0.201 | -0.50 | 0.042 |
| | Substantia nigra | -0.27 | 0.300 | -0.12 | 0.653 | 0.24 | 0.361 |
| | VLPFC | 0.27 | 0.291 | 0.17 | 0.523 | -0.14 | 0.588 |
| | ACC | 0.25 | 0.333 | 0.07 | 0.797 | -0.32 | 0.209 |
| Right | Caudate | -0.41 | 0.101 | -0.21 | 0.414 | 0.31 | 0.231 |
| | Putamen | 0.27 | 0.292 | 0.33 | 0.202 | 0.13 | 0.615 |
| | Globus pallidus | -0.19 | 0.460 | -0.14 | 0.593 | 0.07 | 0.782 |
| | Substantia nigra | -0.32 | 0.222 | -0.33 | 0.217 | -0.01 | 0.969 |
| | VLPFC | 0.23 | 0.381 | 0.15 | 0.563 | -0.10 | 0.702 |
| | ACC | -0.07 | 0.782 | 0.07 | 0.786 | 0.28 | 0.273 |

**Disclaimer/Publisher's Note:** The statements, opinions and data contained in all publications are solely those of the individual author(s) and contributor(s) and not of MDPI and/or the editor(s). MDPI and/or the editor(s) disclaim responsibility for any injury to people or property resulting from any ideas, methods, instructions or products referred to in the content.
